# Reproductive and Child Health Indicators at Primary Health Centers: A Trend of last four years and their comparison with National And State averages

**DOI:** 10.1101/2023.06.14.23291405

**Authors:** Dev Desai, Aparajita Shukla

## Abstract

**Background:** Large amount of data is gathered by Community health centers but remain unanalyzed resulting in poor resource allocation and subsequent poor health outcomes.

**objectives:** to study RCH indicators and compare them with NFHS and SFHS

**Methods:** A record based study for 2017–2021 was carried out among all urban and rural primary health centers of Ahmedabad. RCH indicators were compiled and compared with NFHS and SFHS to draw conclusions about the performance of these centers.

**Results:** in all selected centers, in last 4 years, almost all women who registered pregnancy have come for ANC visits; home deliveries, Health Centre based Cesarean-section, Vasectomy and malnutrition along with Severe Acute Malnutrition has decreased. Female sterilization has remained same. Immunizations has remained satisfactorily. Newborn Mortality Rate, Neonatal Mortality Rate, Infant Mortality Rates, Under-5 Mortality Rate and Maternal Mortality rates all have decreased. This results are similar in both Rural and Urban areas and are along the lines of recent NFHS and SFHS. It was particularly found that Data Management at various levels is faulty and there are mistakes in Data Collection, Data Gathering, Data Analysis and Data preserving. There is a visible difference between how Rural data is managed versus how Urban data is managed.

**Conclusions:** Valuable information is generated which can be utilized by local governing bodies. Better methodology and techniques are now a necessity for quality Data Recording, Management and Safe-keeping. Use of technology can reduce these limitations in the field and improve decision making.

## Introduction

The United Nations declared their 17 Sustainable Development goals (SDGs)[1] in 2016 as per the 2030 sustainable development program. Goal 3 (Good health and well-being) and Goal 5 (gender equality) along with Goal 1 (no poverty) and Goal 16 (peace and justice) are the main leaders fueling an important program called UHC2030 [2] meaning to take action and reach sufficient and sustainable Universal Health Coverage levels across the world.

India has given importance to the Indian Health system and that has led to development of Government based healthcare systems across the country reaching in every village helping hundreds of millions of people in the nation on an annual basis. This system is widely based and dependent on Community Primary Healthcare Center or PHC. A urban Community healthcare center is called U-PHC or UHC meaning an Urban Health Center while a PHC in rural areas is called a R-CHC or P-CHC or PHC only, meaning a Primary healthcare center. This infrastructure has enabled the Indian government and medical system to reach a depth of populations like never before. This completely free and yet, efficient medical health care system has led to decrease in many diseases’ prevalence, helping us eradicate many deadly diseases (e.g. Polio), assisting us to reach people in the rural and urban, approachable or non-approachable, poor or rich to cater to their needs, address it and helping them at our full potency.

In rural settings, PHCs work as a beacon of healthcare to the people but also work as a Referral Center to send patients to Secondary healthcare centers Based in Taluka capital and to Tertiary healthcare centers based in District Capitals and cities.

These efforts are highly socially accountable in nature and hence have to be evaluated periodically. For evaluation purposes, every CHC gathers data of patients or rather the data about the benefits given to the population by each CHC to keep a record of its functioning as well as budgeting and future references.

One of the main goals in UHC2030 is healthcare to women and children, especially in the regions and areas where they have been overlooked for years. India has been working towards the same goal of healthcare to every child and women, especially women in reproductive age and who are pregnant.

As mentioned above, CHCs gather what is known as RCH indicators (Reproductive and child healthcare)[3] data which are usually analyzed by the authorities for future references as well as to check proper functioning and efficiency of the CHC. Reproductive indicators are the best way for a country to assess Reproductive health of the citizens. Many of the health problems of Indian women are related to or exacerbated by high levels of fertility. The poor health of Indian women is a concern on both national and individual levels. It affects the children who will be India’s next generations of citizens and workers[4]

## Review of literature

Indian government has introduced many Schemes and incentives like Free national children immunization, Ayushyaman Bharat etc. for the benefit of women and children to bring better Reproductive and Child health into existence and mainly to decrease morbidity, complications and mortality in children and mothers. These steps have indeed played a visible role in the healthcare system and healthcare delivered to Indians and the indicators which are used to keep track of total aid that has been given and its impact on the community are increasing only indicating we are on the right track. These analytical indicators hold a special importance as they show what is happening in the whole country and also demonstrate what is happening in a single PHC or a small remote village.

Proper independent analysis of these indicators should also be carried out to know the true numbers. This will help in understanding the need of redistribution and reallocation of resources as to reach more and more citizens.

U-PHC (UHC) report their data to Municipal corporations while PHCs report it to District CDHO (Chief District Health Officer). This data eventually ends up at the state government for the formation of SFHS (State Family Health Survey)[5] and then it is delivered to the central government for formation of NFHS (National Family Health Survey)[6]. Date is tried to be evaluated at every level and needs are assessed at every level as per their jurisdiction.

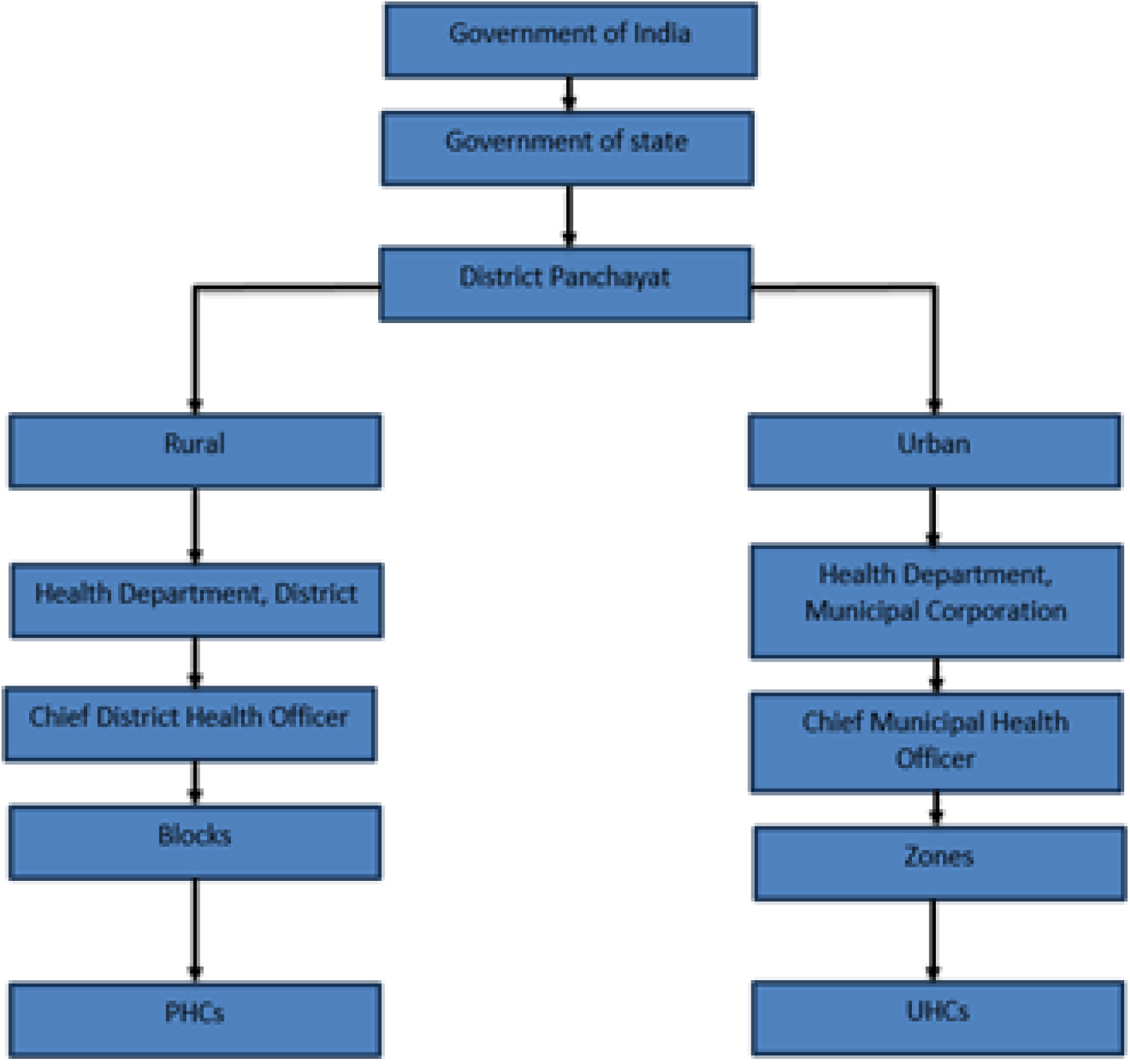

**Aims and Objectives:**

● To study RCH indicators of concerned PHC and UPHC
● To compare this data with NFHS and SFHS
● To compare indicators of UPHC and PHC
● To study the quality of work done by individual PHCs and to recommend if resource redistribution is needed or not.

## Material and Methods

This study is a record based study undertaking PHC and UPHC as Study area.

Inclusion criteria:

⍰ Primary health center both Rural and Urban.
  ○ Whose data is available and trustable from respective authorities with their consent.

Exclusion criteria:

⍰ Only primary health centers are considered here
⍰ Secondary and tertiary health care centers (including hospitals) are not considered due to unavailability of the data or inconsistency of the recorded data
⍰ Private hospital sector is not considered here due to unavailability of the data and inconsistency of the recorded data. Frankly, due to inconsistency, there is no way to tell whether at some places private hospital data is added or not.

Recently, NFHS-5 state data sheet [7] was released by the Indian government, hence NFHS-5 Gujarat[8] data is considered in this report.

Even though the NFHS-5 state data sheet has been released, total pooling of all state data is not yet, hence for comparison with National Averages, NFHS-4 India [9] is considered.

For the purpose of the study different Zones of Urban Area (Ahmedabad Municipal Corporation is divided in 7 zones and UHCs are distributed among these 7 zones) and different blocks of rural Area (Rural/ Ahmedabad District (excluding the urban area) is divided in 9 blocks)) are compared. Also, Comparison is made with State SFHS and National NFHS data wherever possible of the selected RCH indicators for the report.

## Observations and Results

Data from Ahmedabad Municipal Corporation was requested and data of every UCHC was recorded. Similarly, Ahmedabad District Panchayat was requested under which data from Rural PHCs was gained and recorded. Data was pulled in Zones (in UCHCs) and Blocks (in PHCs) for a better understanding and comparative analysis.

## Discussion

As it can be seen, different parameters are compared between different zones and they are also compared with different blocks that come under the geographic definition of Ahmedabad district and municipality.

Zones are part of the Ahmedabad city proper in Ahmedabad District which is controlled by Ahmedabad Municipal Corporation whereas Blocks come under the supervision of Ahmedabad District Panchayat.

### 1) Total number of pregnant women registered for ANC

As it can be seen here in Figure 1.1 Central, North West and South West zones show lesser numbers of registered pregnancies compared to other zones. It can also be seen that in the years 20-21, registered pregnancies in all zones is less than what was noted in previous years in respective zones.

In Figure 1.2, It can be seen that there is a sharp decrease in registered pregnancies in Daskroi whereas an increase is visible in Detroj and Dholka blocks. In most of the blocks the numbers of registered pregnancies remain the same with time. This shows that there is some misconduct in data collection, management and reporting.

**Figure 1.1.**
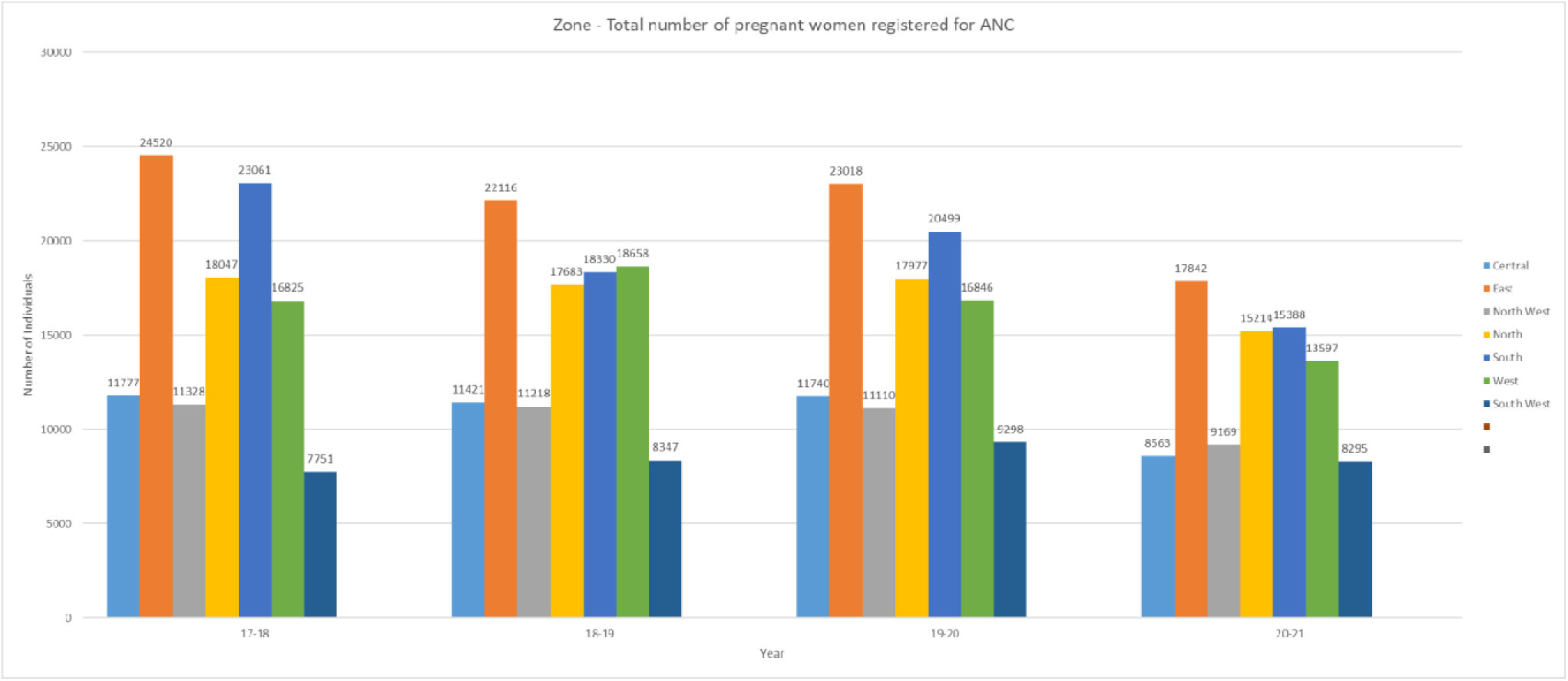

**Figure 1.2.**
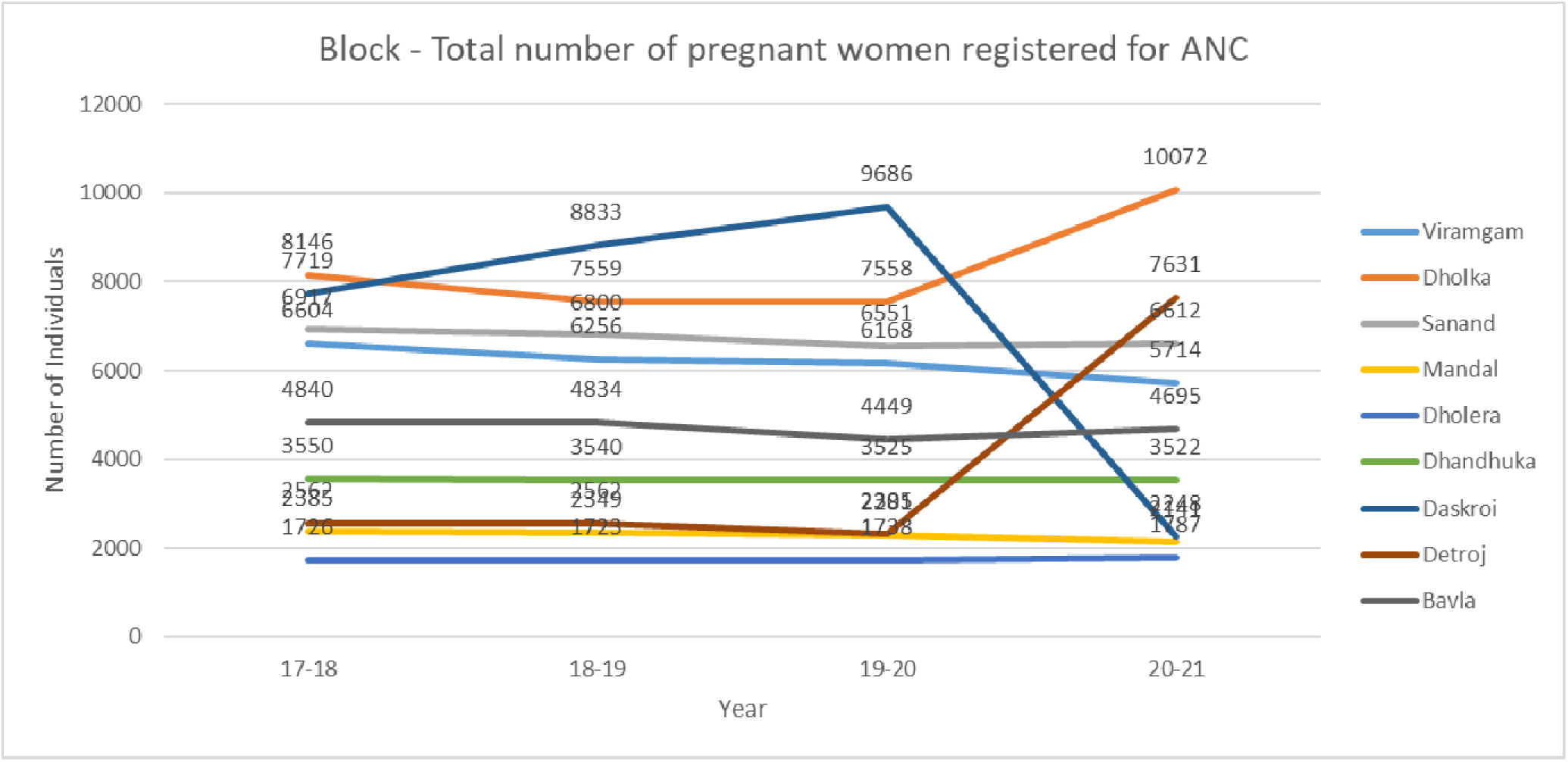

### 2) Number of PW received 4 or more ANC check ups

As it can be seen here in Figure 2.1, registered pregnant women showing up for minimum required 4 ANC visits are highest in East zone and lowest in South Zone (Maybe because East zone has the highest % of low socio-economic class citizens). Comparing it with Figure 1.1, we can see that not all women who registered are showing up for ANC visits. **All women who get pregnant are registered even though the pregnancy is diagnosed in the private sector or public setting. But track of whether ANC visits are done or not is only done at women coming to public settings. Hence, valuable information is lost in the process.**

In Figure 2.2, It can be seen that there is a sharp decrease in ANC visits in Daskroi whereas an increase is visible in Detroj and Dholka blocks. In most of the blocks the numbers of ANC Visits remain the same with time. This can be guessed from figure 1.2 and it is evident that trend of women completing 4 ANC visits is based on and hence similar to Registered pregnancies.

Figure 2.3 denoted that **as Rural places have less tertiary care hospitals, more ANC checkups are done at public settings hence more % of pregnant women can be seen in all years compared to Zonal (urban) level with high density of private hospitals.** Data recorded in Gujarat shows lesser number of ANC visits in rural blocks than in Ahmedabad but higher number of urban visits than in zonal Ahmedabad city. **The reason behind this could be that as private sector hospitals in Gujarat are concentrated in Ahmedabad, other areas classified as urban have less private hospitals so people living in these areas also go to public settings.** Total average of India as per NFHS-4 seems lower than Gujarat in NFHS-5 meaning more pregnant women % of pregnant women in Gujarat are getting ANC visits then in India.

**Figure 2.1.**
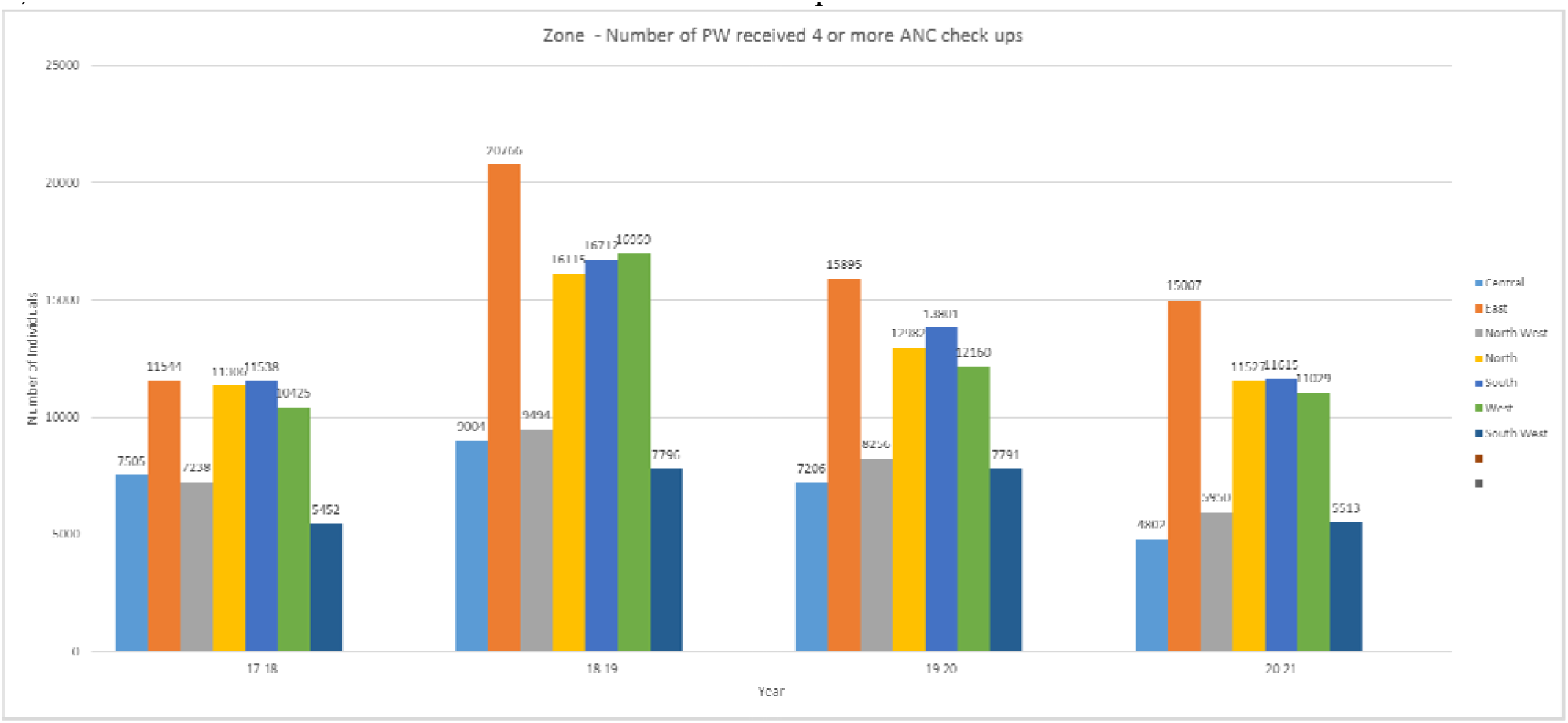

**Figure 2.2.**
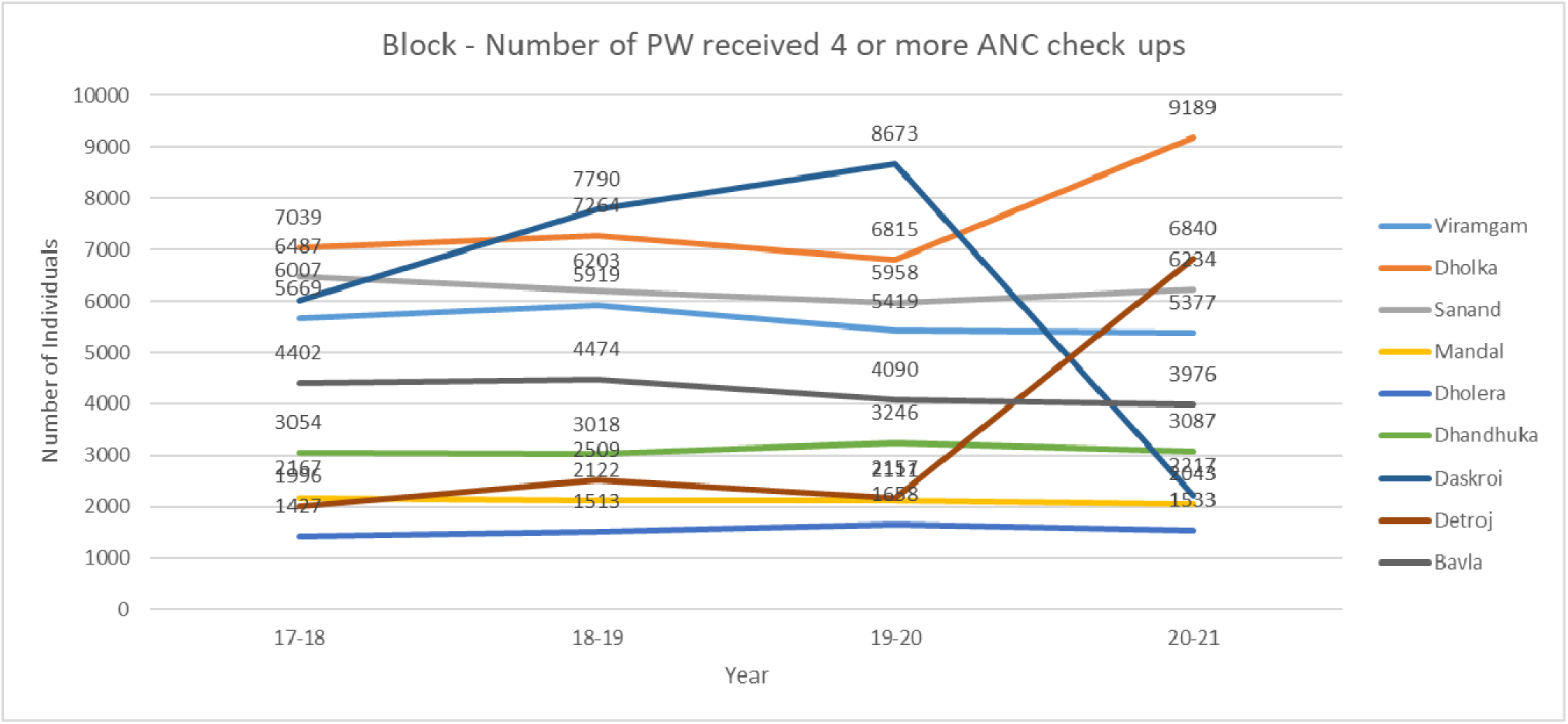

**Figure 2.3.**
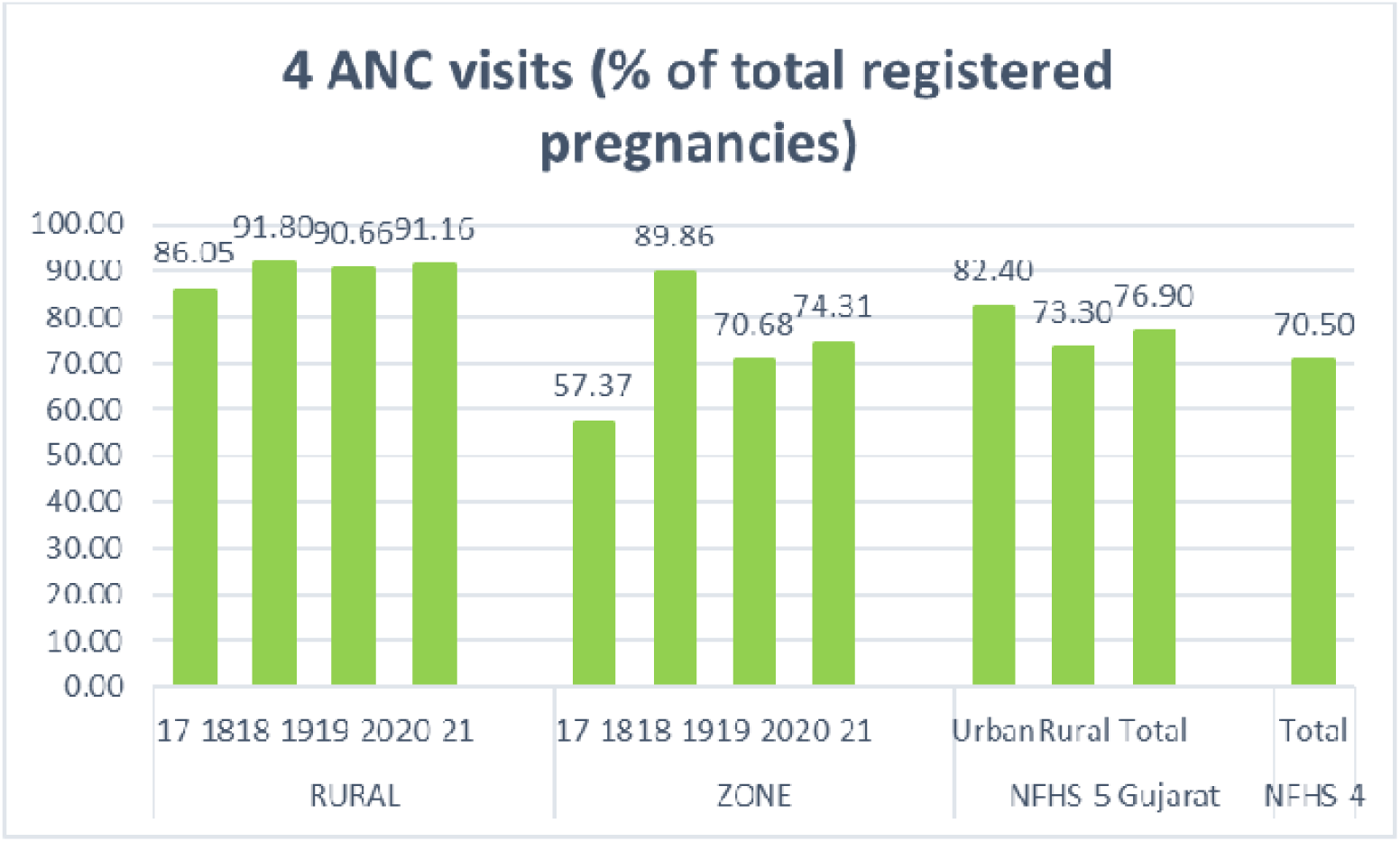

### 3) Number of Home Deliveries attended by Skilled Birth Attendant(SBA) (Doctor/Nurse/ANM)

From Figure 3.1 and 3.2 it is visible that the total number of SBA attended pregnancies are too low. Highest amount of these home deliveries can be seen from Central zone then east and North zone (Maybe because East zone and then North zone has the highest % of low socio-economic class citizens). Talking about Ahmedabad rural it is very low, almost 0. **The probable reason behind this as per us is as all hospitals were assigned to handle covid patients, there was a shortage of hospitals and public setting centers to aid with delivery. Also due to the pandemic there was a fear of Covid-29 which could have led to people preferring Home deliveries instead of institutional delivery.**

Figure 3.3 shows that in Ahmedabad rural, there is very low SBA home deliveries suggesting a strong referral system and rural PHC Centre for deliveries or maybe under reporting of SBA deliveries. Whereas areas considered in Urban have a higher SBA % a**s live births in tertiary care hospitals and private settings are not counted in RCH data taken by the Municipality while preparing the information.**

Under the RCH program there is a provision for a maternal **and child tracking system** in the national program but at zonal and block level that data is not aligned properly and there is total data mismatching between UHC data and true number hence it becomes challenging to compare UHC Ahmedabad data with Indian averages. Also the data mentioned here does not say anything about the procedures and outcomes in a private setting. It is also not clear whether that data is counted here or not.

**Figure 3.1.**
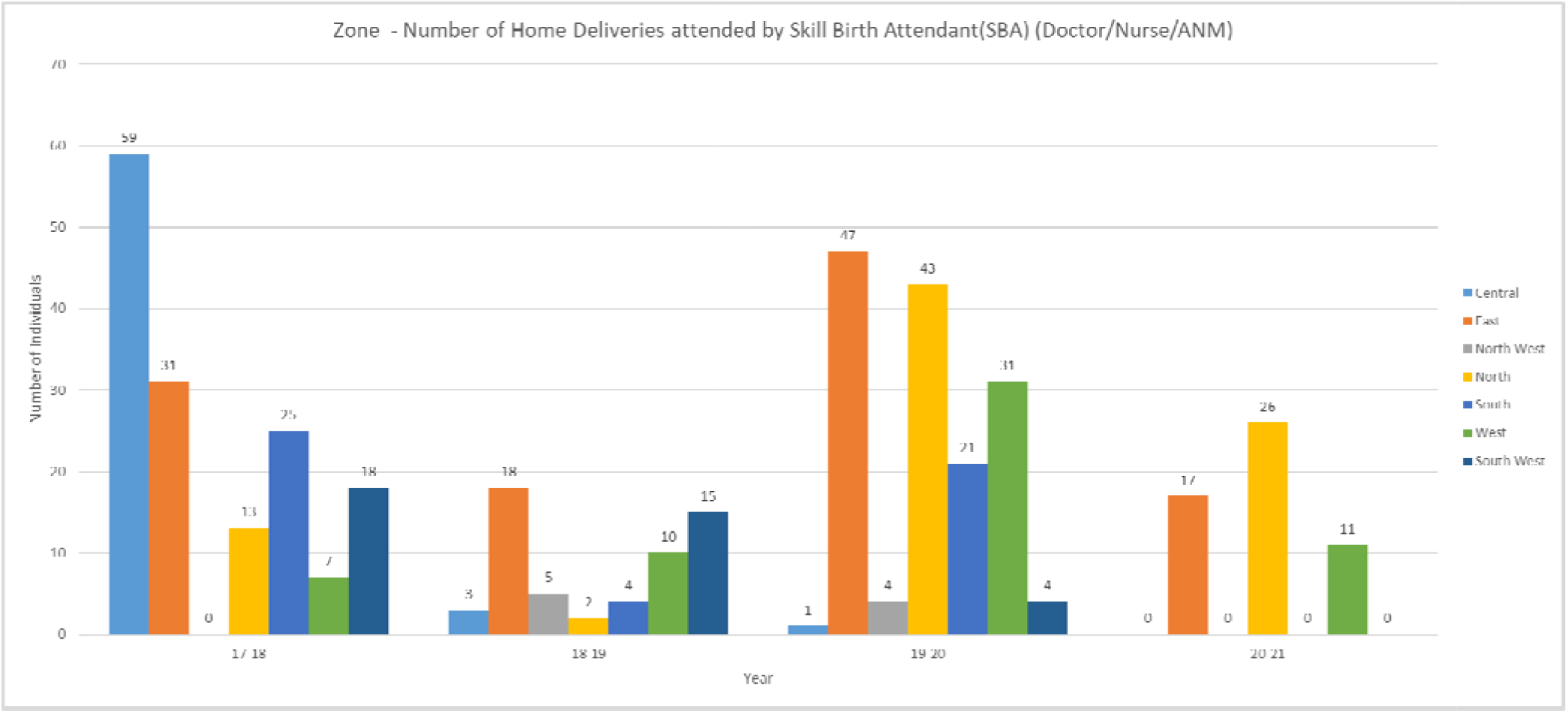

**Figure 3.2.**
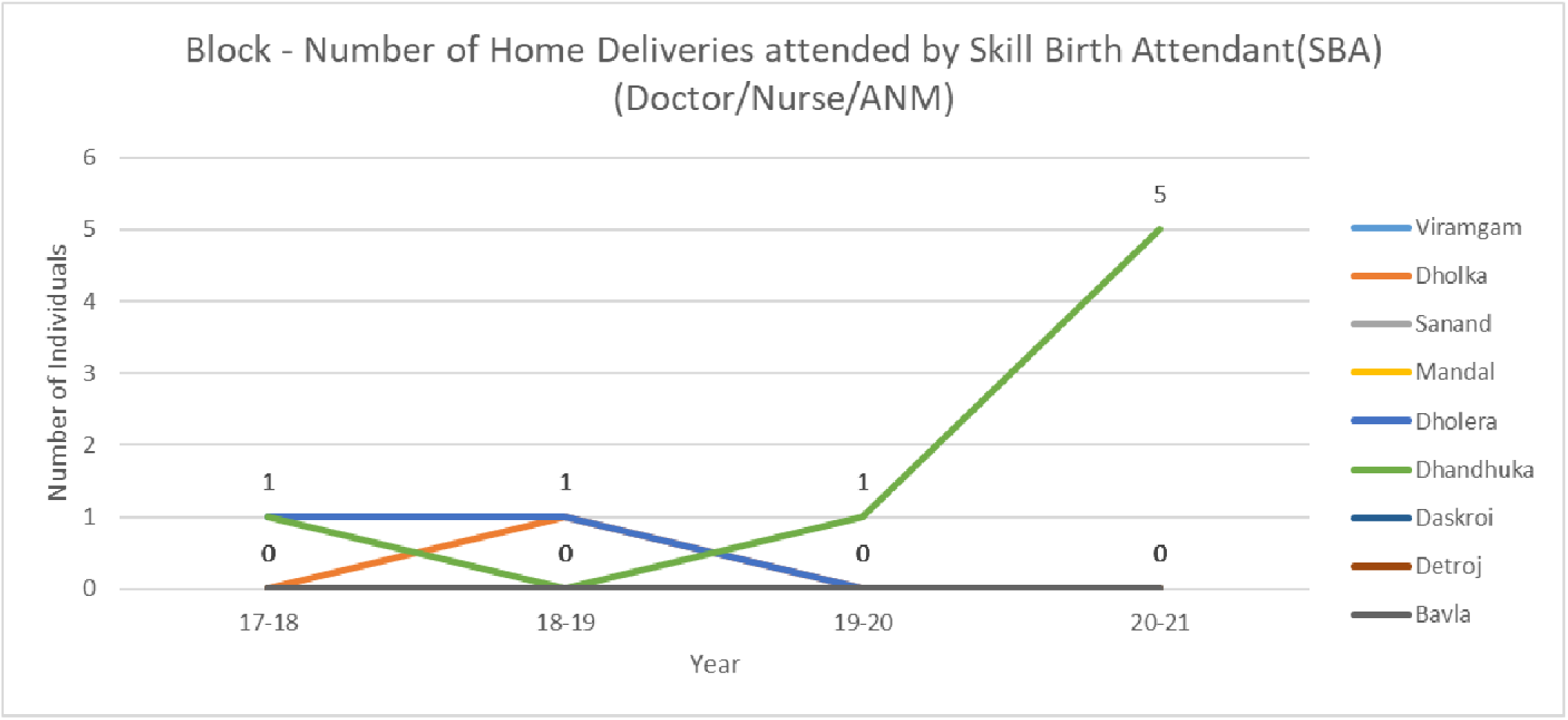

**Figure 3.3.**
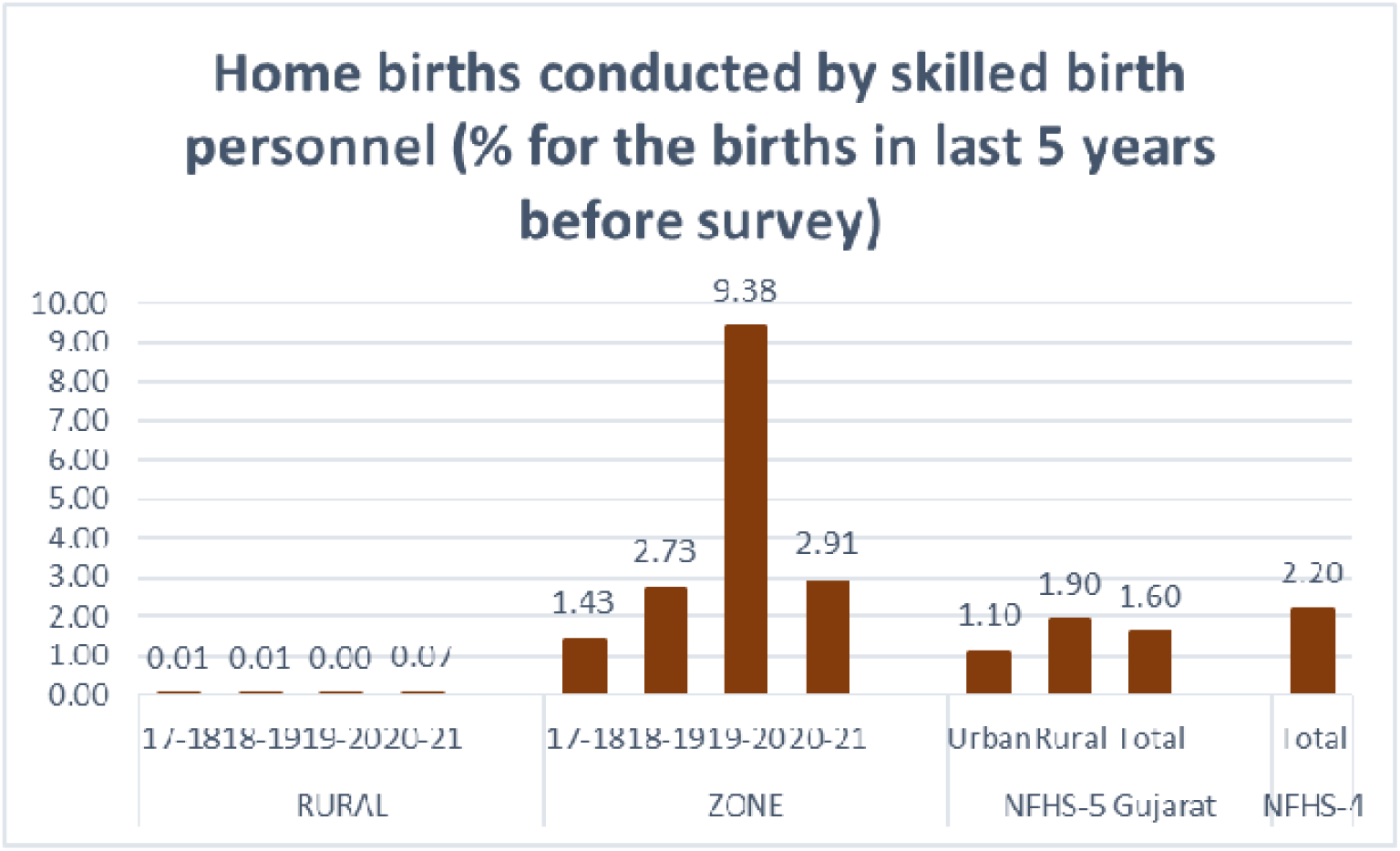

### 4) Number of Home Deliveries attended by Non SBA (Trained Birth Attendant(TBA) /Relatives/etc.)

Figures 4.1 and 4.2 Suggests that Non-SBA deliveries have had reduced in year 19-20 in urban Ahmedabad which has risen in the south zone while it has decreased to 0 in Rural Ahmedabad.

There also seems to be some confusion and comparison in data entry where SBA might have been understood as TBA and TBA as SBA.

**Figure 4.1.**
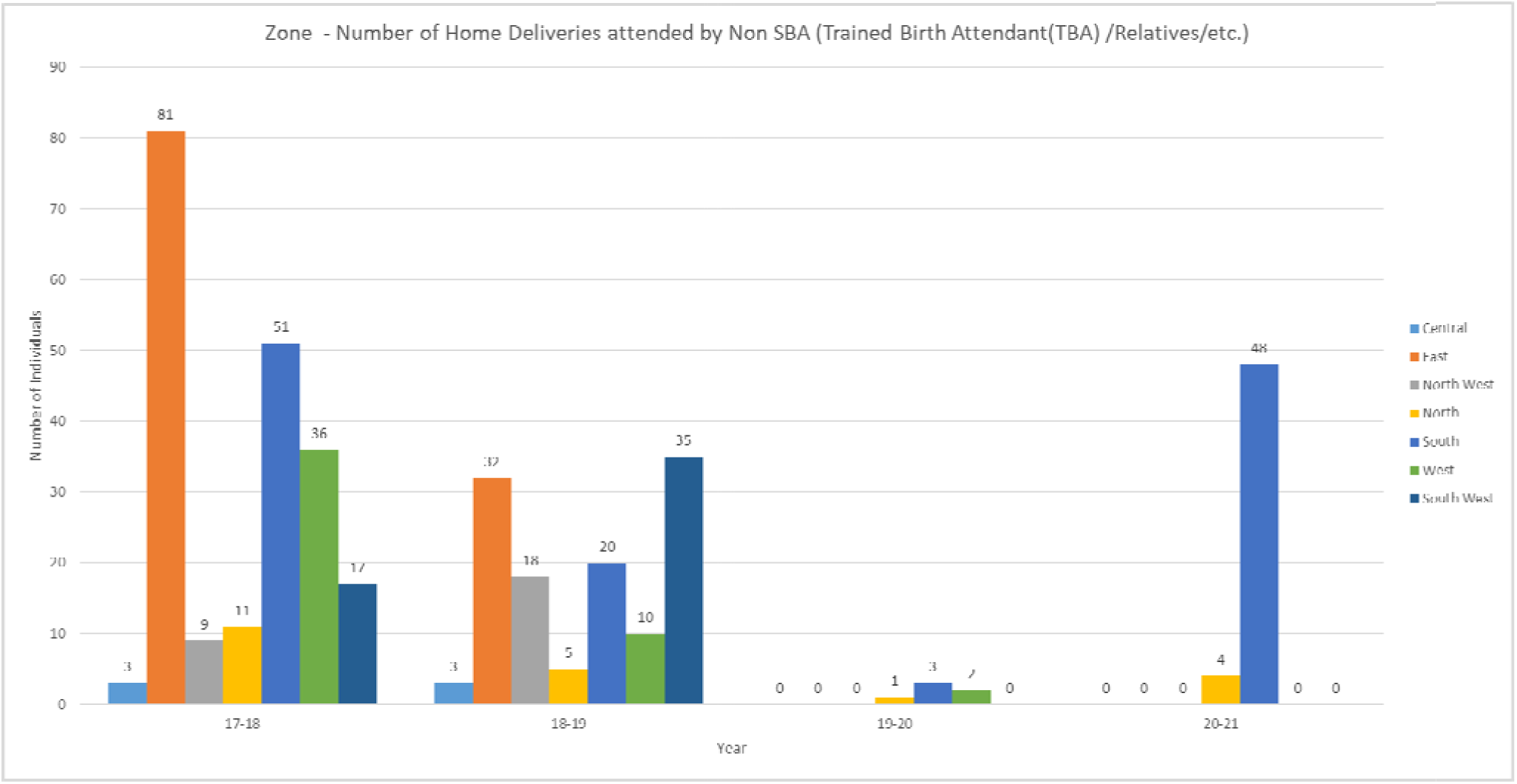

**Figure 4.2.**
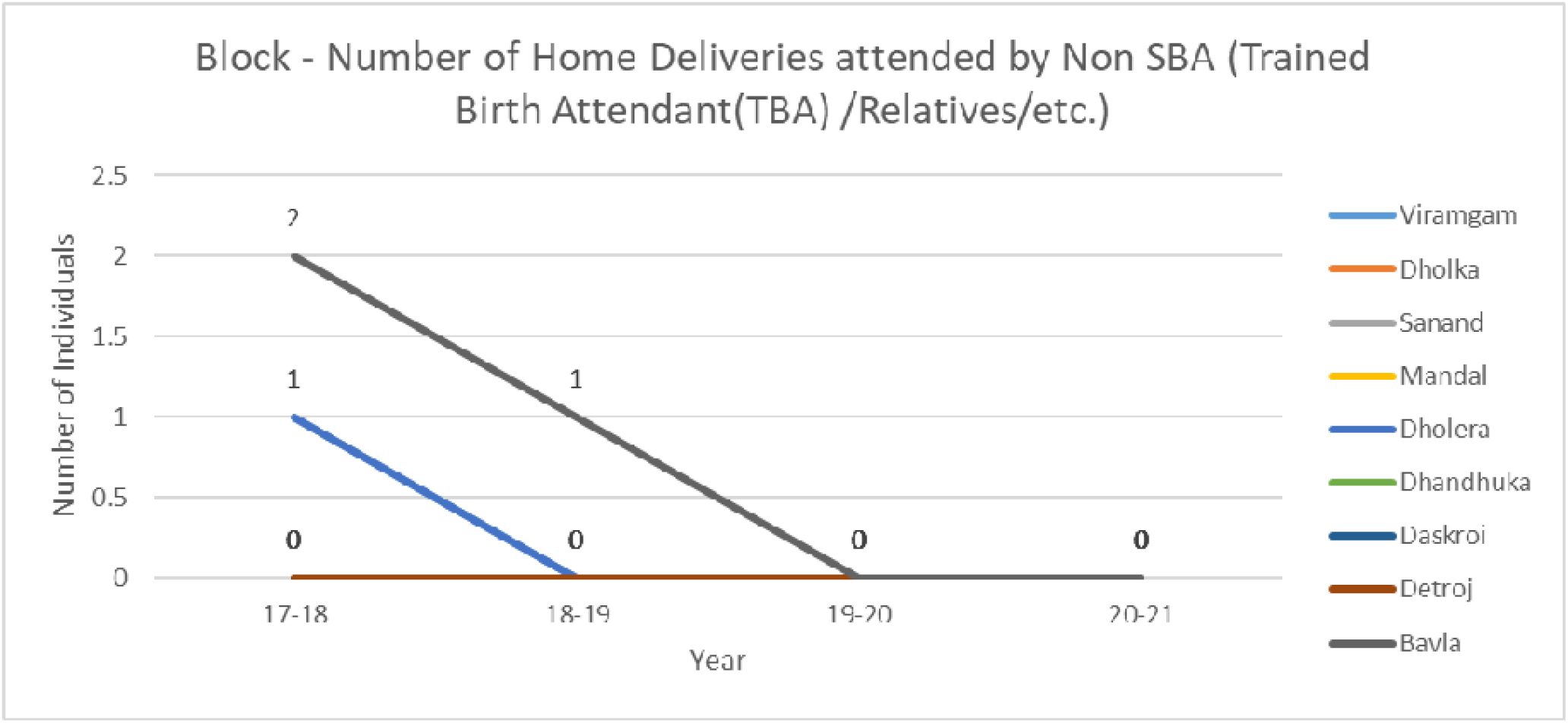

### 5) Number of Institutional Deliveries conducted (Including C-Sections)

Figure 5.1 shows that the total trend of institutional deliveries has gone down in urban Ahmedabad with years passing but as shown in figure 5.2, an increase is visible in institutional deliveries till 19-20 which shows a significant drop in year 20-21.

Figure 5.3 gives enough evidence as to why this type of study is required as huge difference is visible. Rural Ahmedabad shows a very high number of institutional deliveries giving a hint that deliveries in **private hospitals in rural areas was also accounted as institutional deliveries.** The increase in Urban institutional deliveries % is not understandable. **Data available as a part of NFHS-4 and NFHS-5 Gujarat shows a very different side and hence, there is disparity in the data fields which cannot be understood.**

**Figure 5.1.**
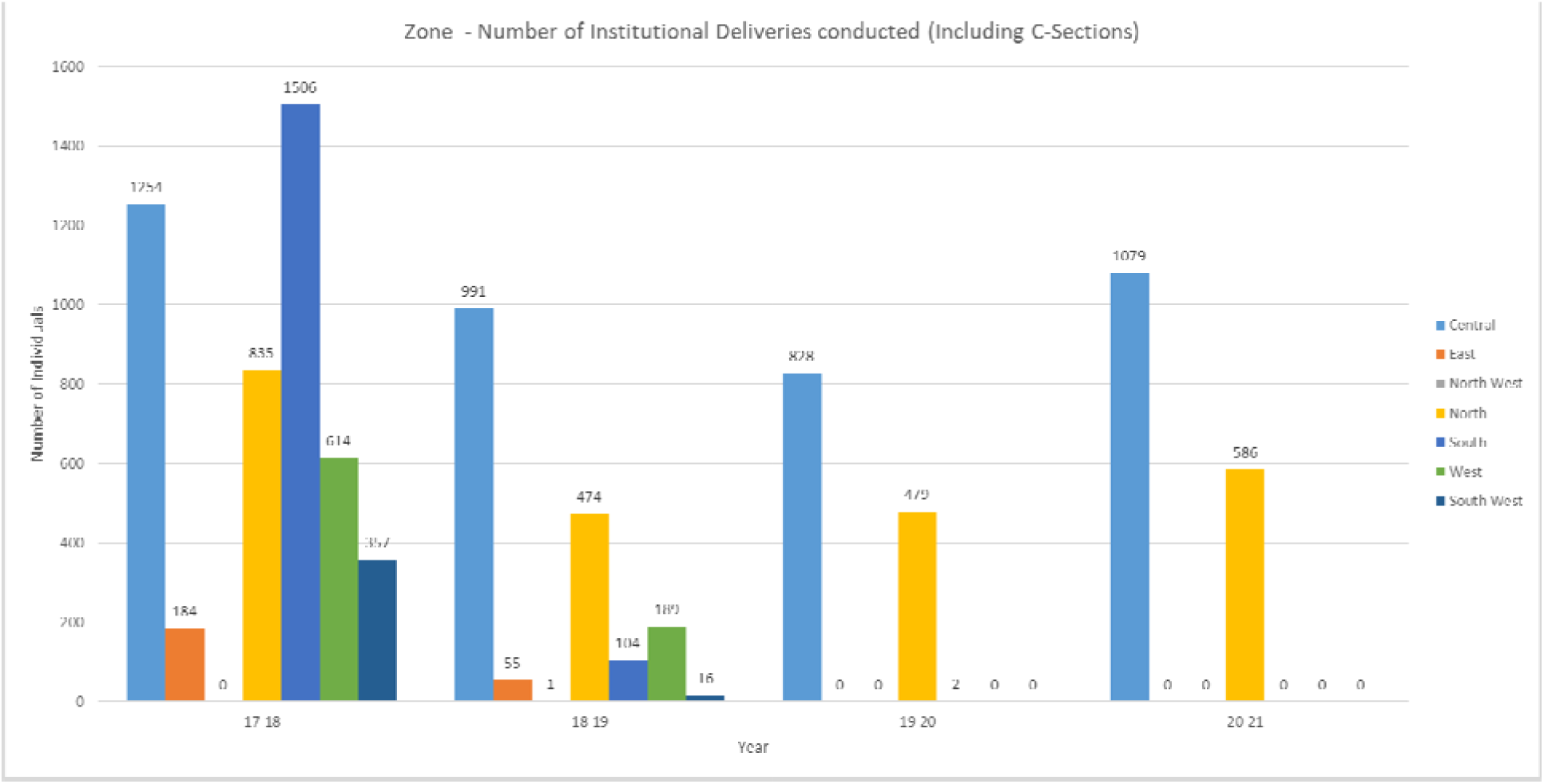

**Figure 5.2.**
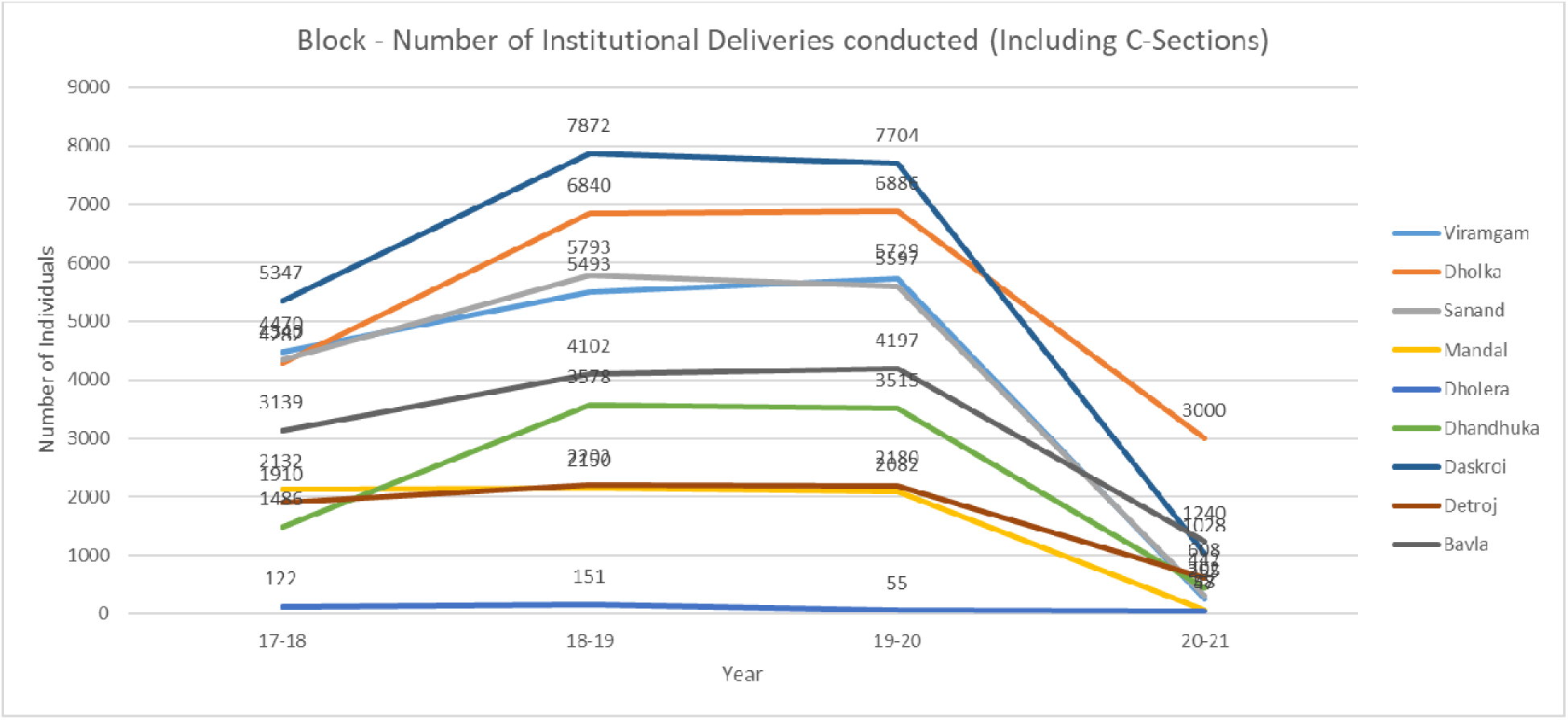

**Figure 5.3.**
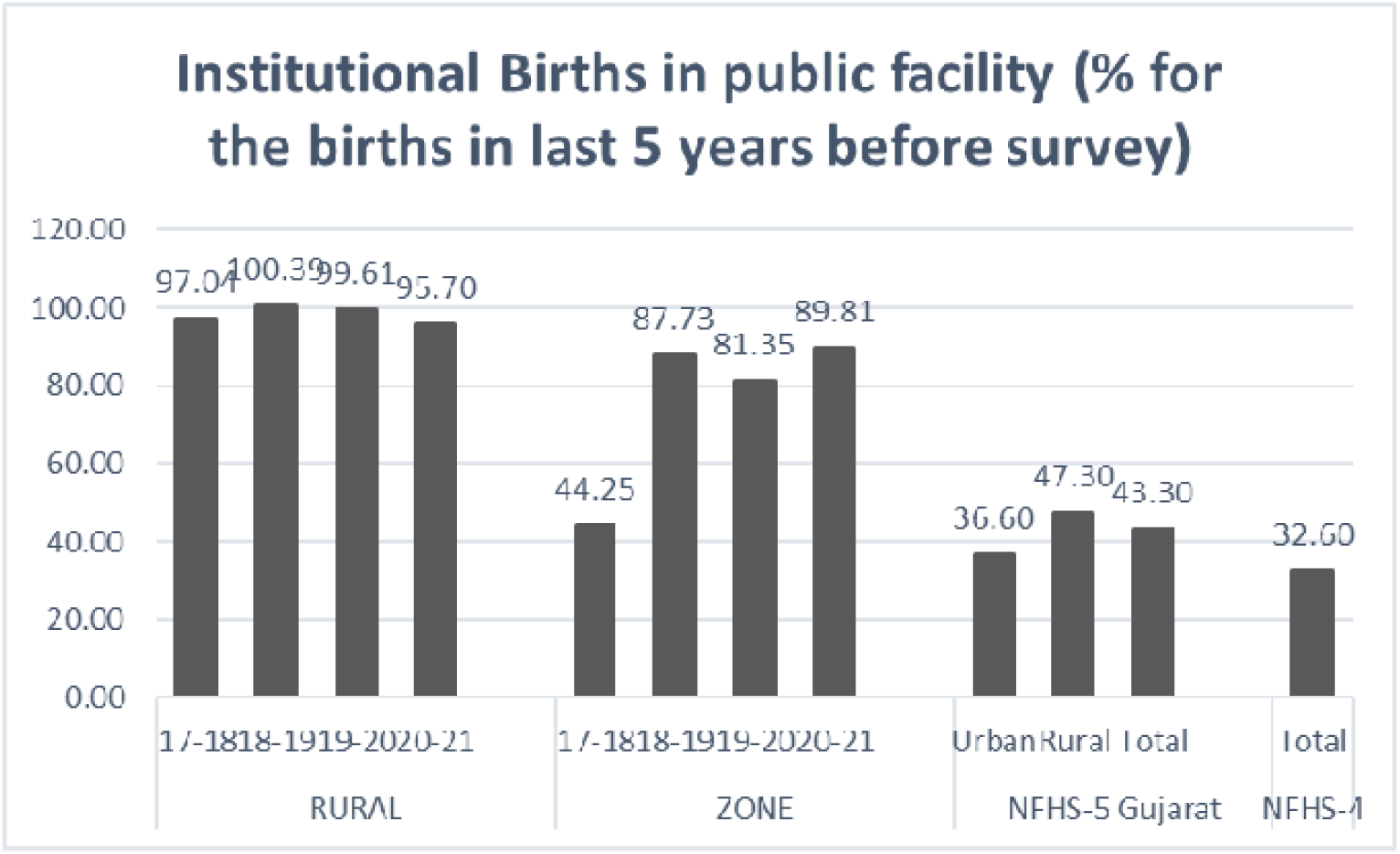

### 6) Total C-Section deliveries performed

As described in figure 6.1, a trend can be seen over the year where the total number of institutional CS deliveries are decreasing to 0 in the year 20-21.

In rural Ahmedabad as per the figure 6.2, a constant level of institutional CS has decreased significantly.

Figure 6.3 helps us understand that in the total live births registered, a range of 15-20% are born with the help of CS. This average % match with Gujarat data and NFHS-4 India.

**Figure 6.1.**
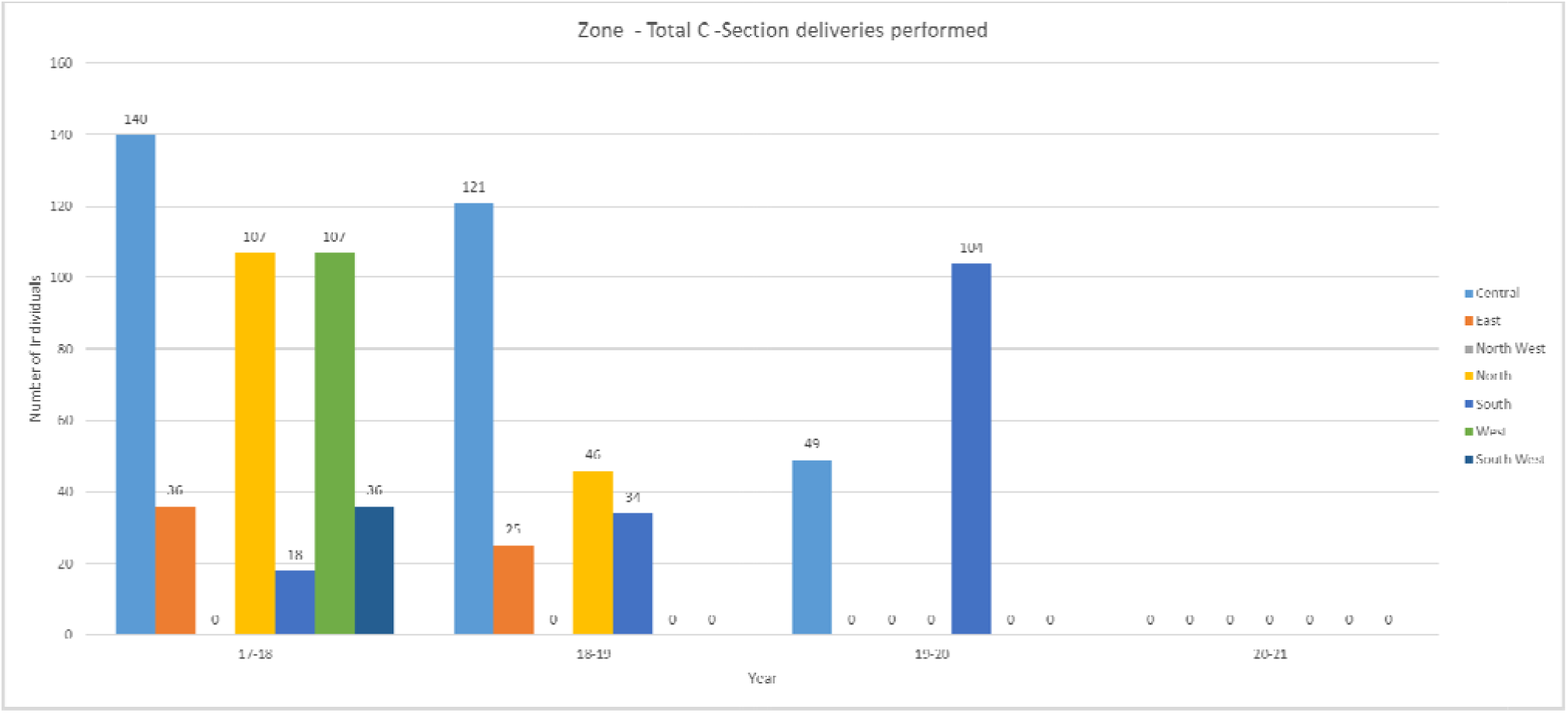

**Figure 6.2.**
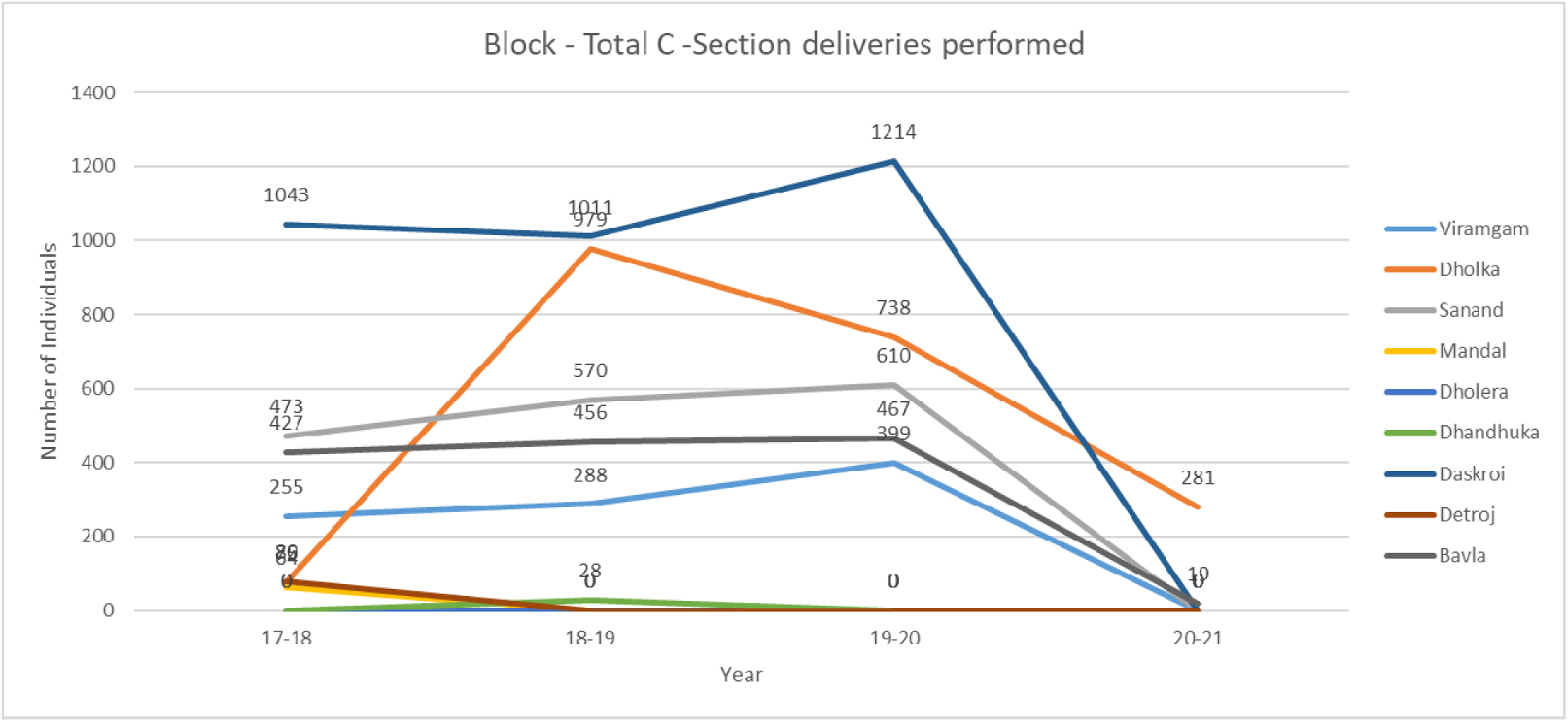

**Figure 6.3.**
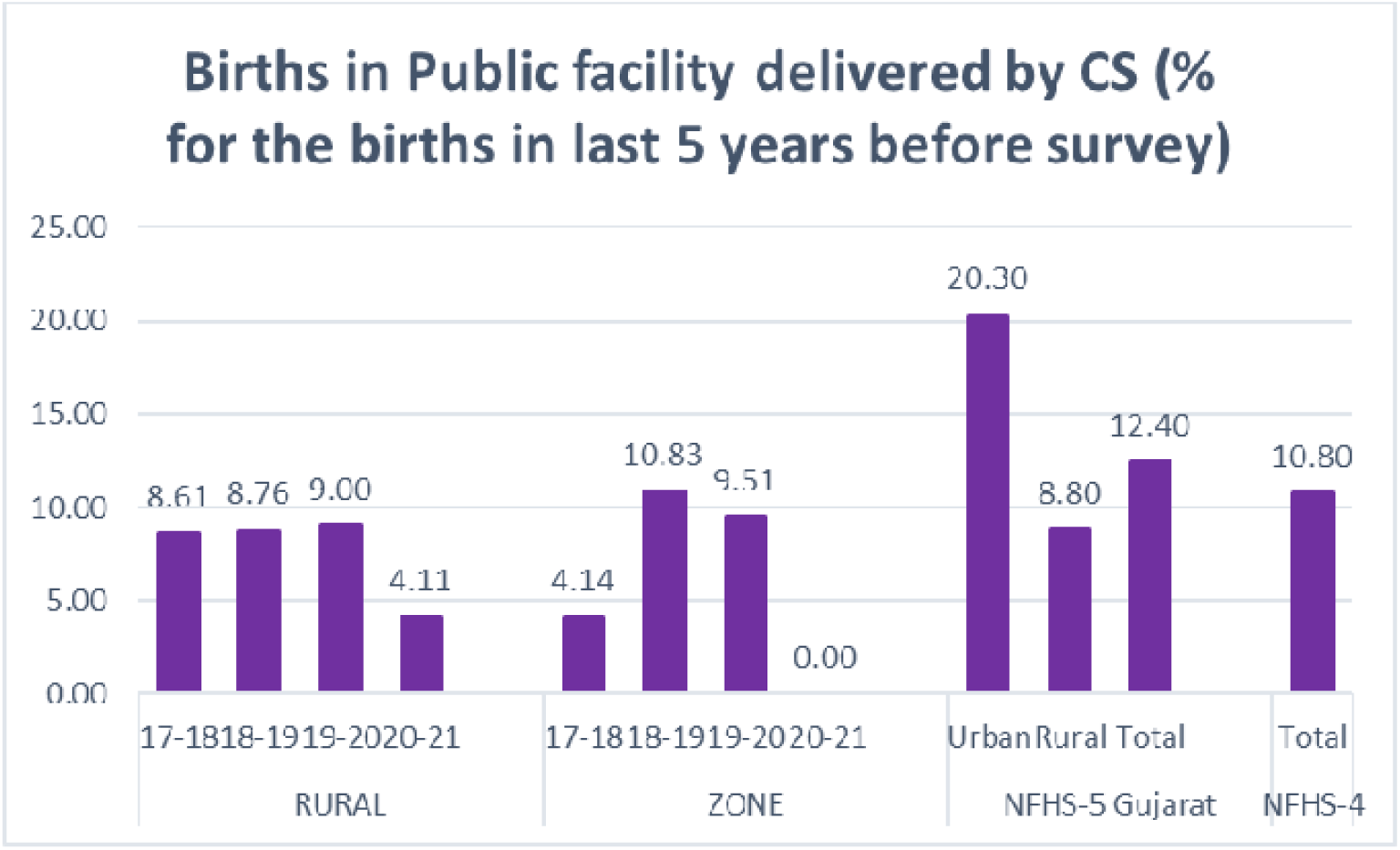

### 7) Live births

Figure 7.1 and 7.2 try to show the live births at the institute in Ahmedabad which show a decrease. A constant decrease in the Urban Ahmedabad whereas a peak at year 19-20 and then decreasing significantly in 20-21

The main reason behind this values is that private hospital data is not recorded in these and hence, even registered pregnant women who was showing up for ANC visits here goes to have her child delivered at private hospital which is why, the numbers of live births are so low in compared too registered pregnant women or women who completed 4 ANC visits.

This shows that there is misconduct in data assessment due to mishandling of data, faulty and false entries, poor data storage and management.

**Figure 7.1.**
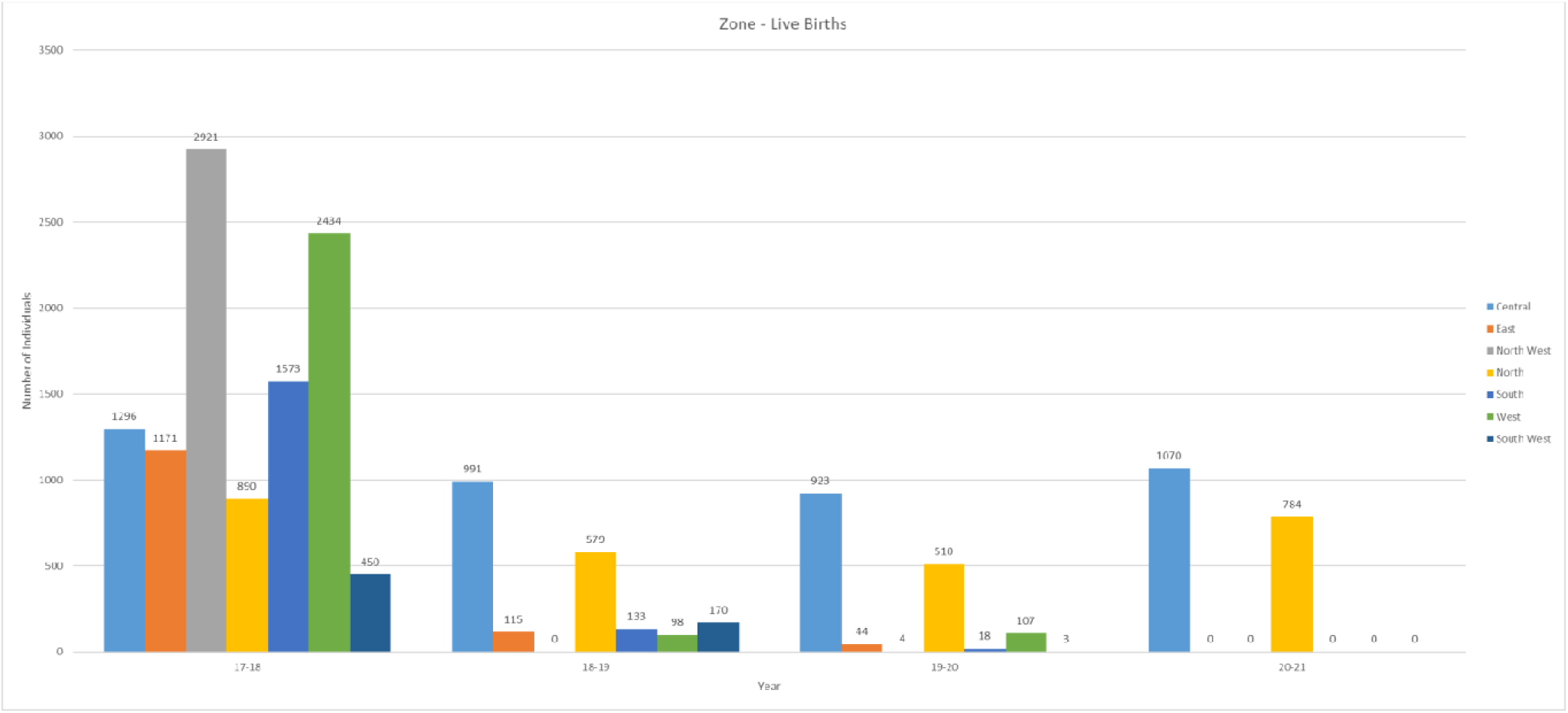

**Figure 7.2.**
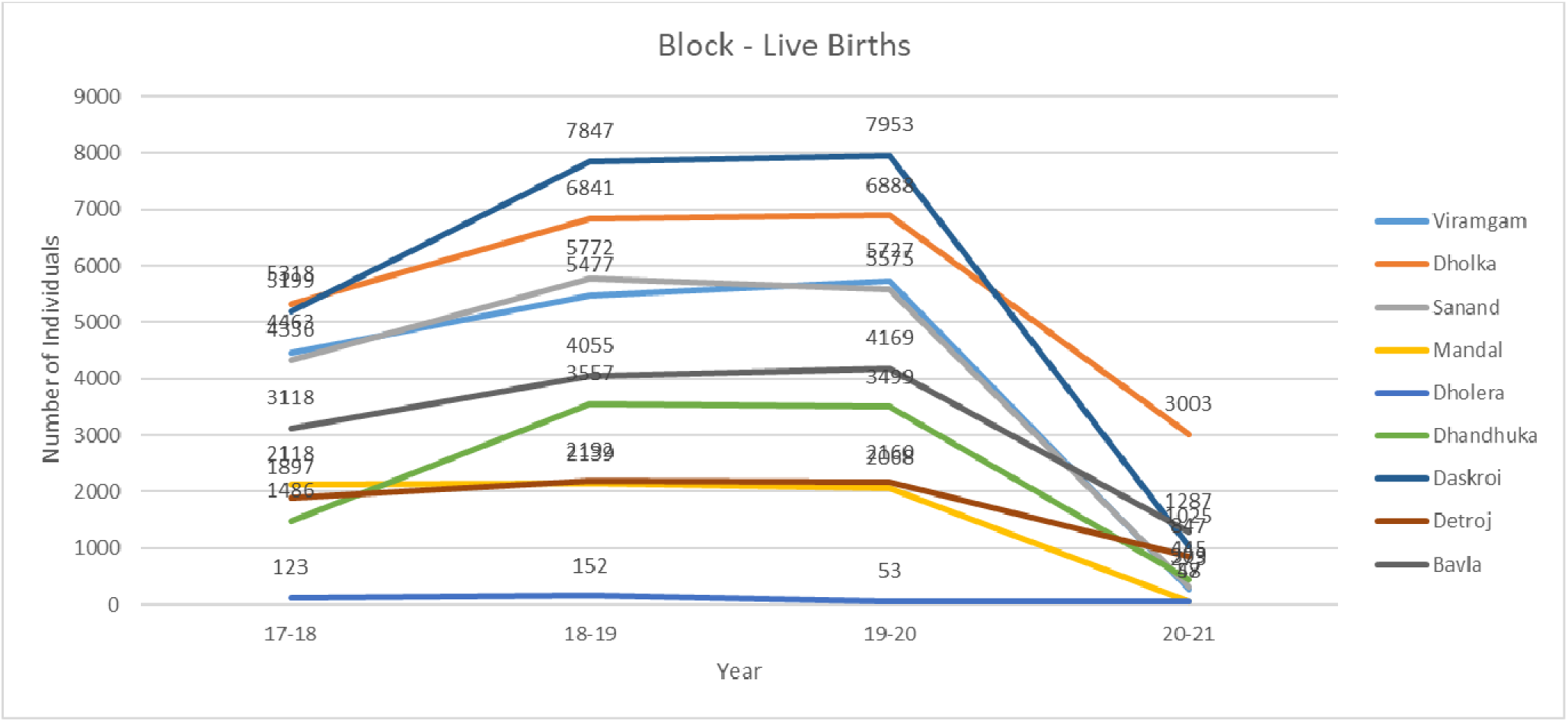

### 8) Number of Newborns breastfed within 1 hour of birth

Figure 8.1 denotes that recording of women breastfeeding in the first hour was kept way better in urban Ahmedabad and the system of recording has worsened with passing time. Comparing it with Figure 7.1, we can see that there are not many correlatable points between the number of live births recorded and the number of children breastfed in the first hour after their birth.

Figure 8.2 helps us understand that record is kept better in rural Ahmedabad then in Urban Ahmedabad and it can actually be compared with the number of live births in rural setting. Yet as seen in all other previous parameters, an increase till 19-20 year can be appreciated which falls significantly in 20-21.

Figure 8.3 helps us understand that Rural Ahmedabad and Urban Ahmedabad are way better in breastfeeding the newborn in the first hour then when considering whole Gujarat as in NFHS-5 or India in NFHS-4. **In rural Ahmedabad, about 99% of newborn were and are breastfed in the first.** When Urban Ahmedabad is compared, it can be seen that there are gross mistakes in calculating the amount as in some years, gross babies breastfed in the first hour is even higher than the total number of live births.

These shows that there is misconduct in data assessment due to mishandling of data, faulty and false entries, poor data storage and management.

**Figure 8.1.**
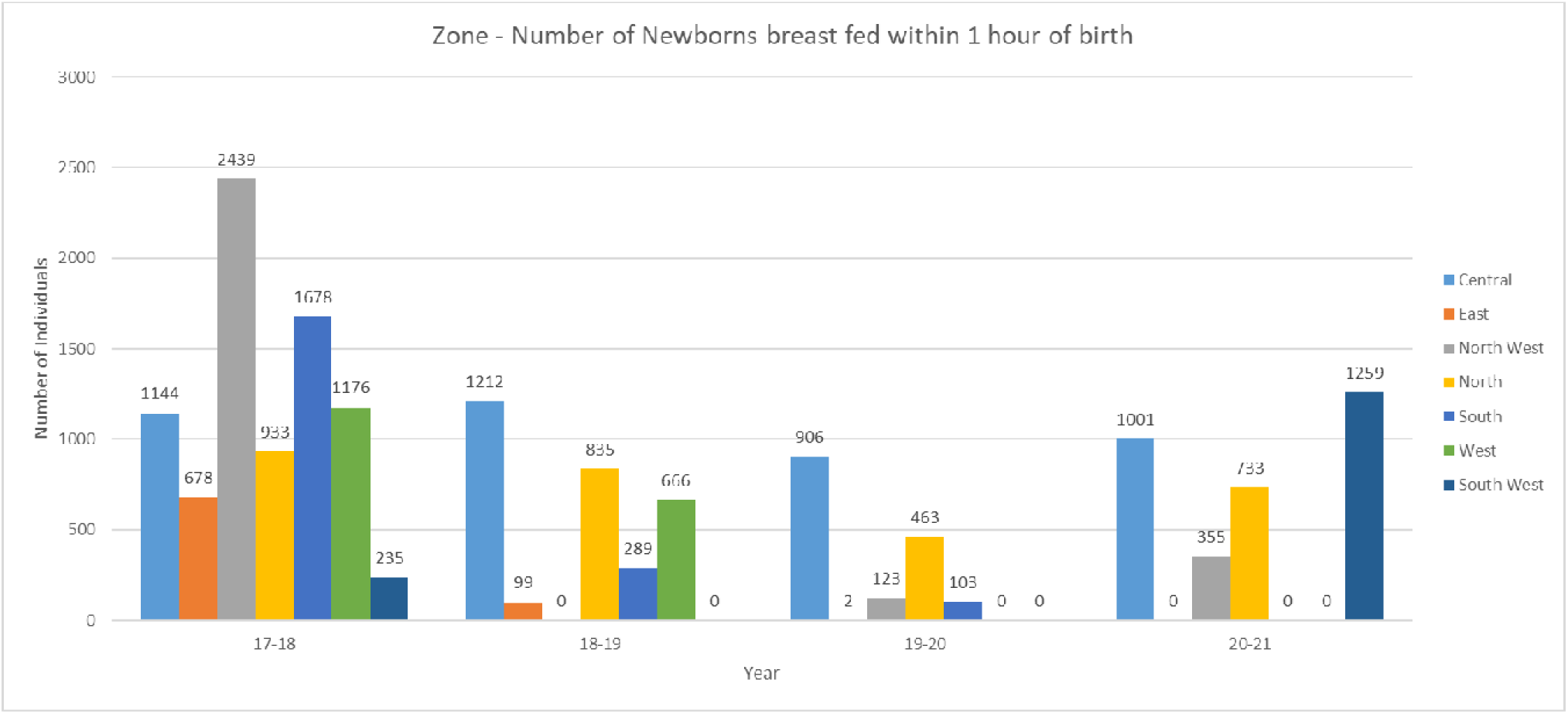

**Figure 8.2.**
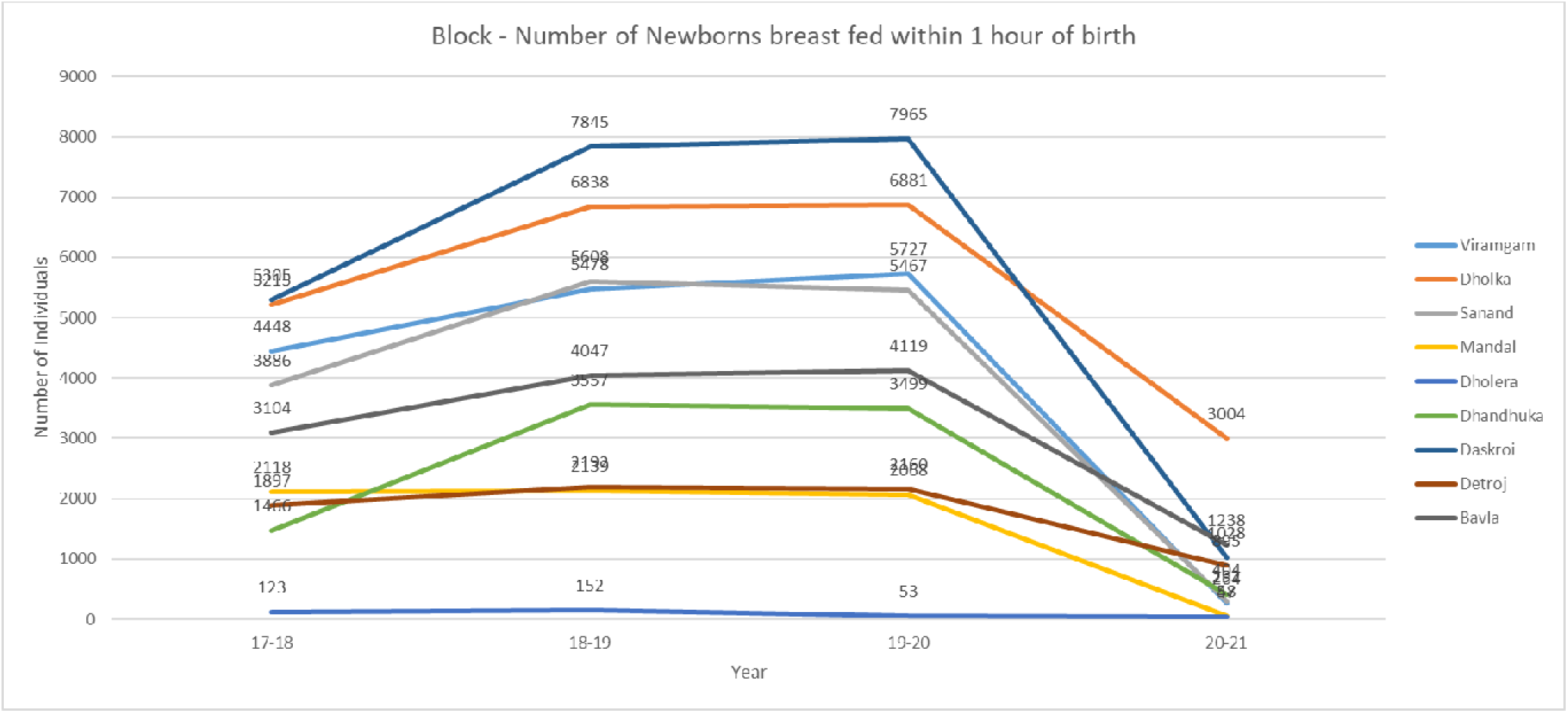

**Figure 8.3.**
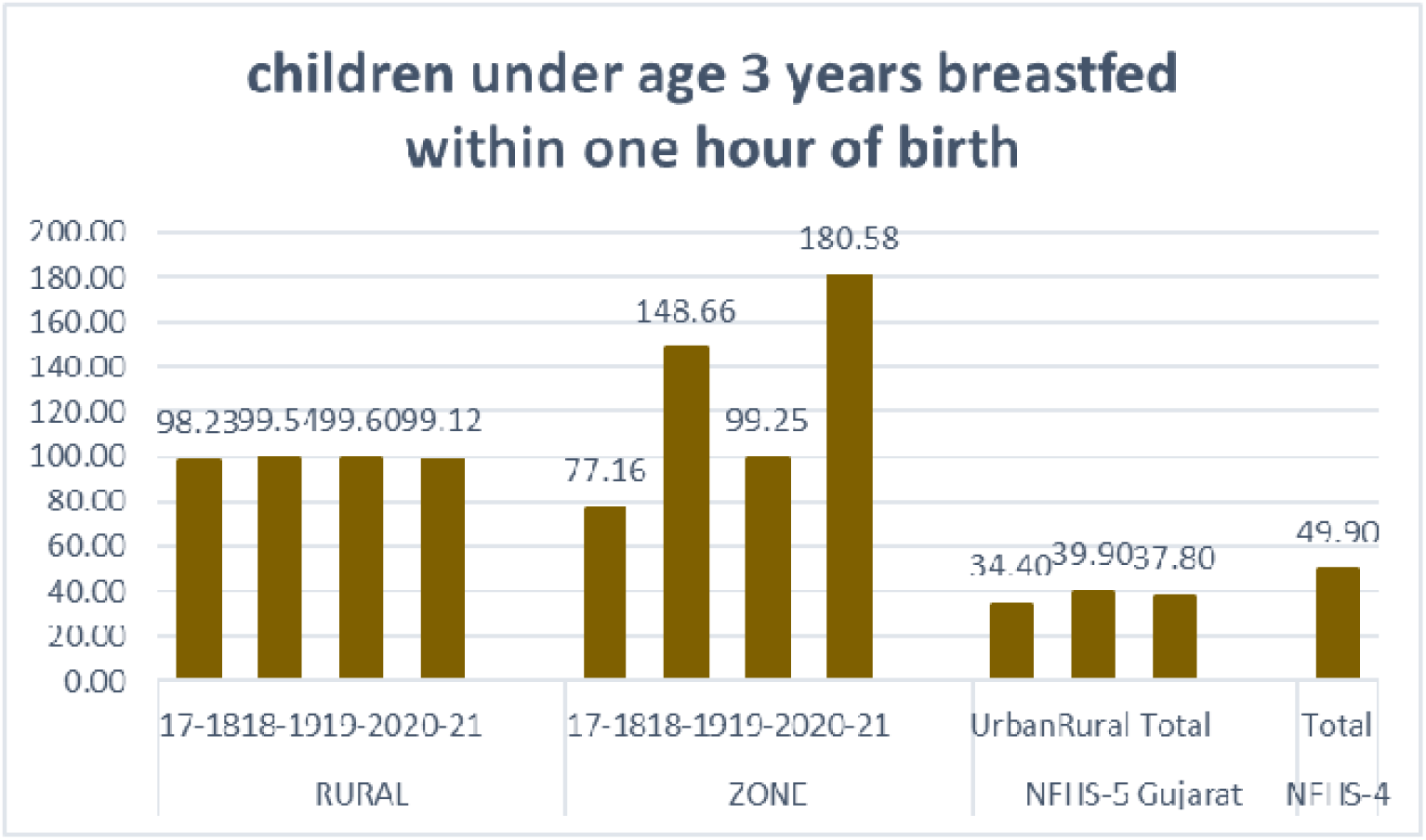

### 9) Number of Non Scalpel Vasectomy (NSV) / Conventional Vasectomy conducted

Figure 9.1 helps us understand that the total trend of having a registered Vasectomy has decreased over the years in urban Ahmedabad. The highest numbers are still seen in the East zone while the North West zone shows the lowest number of NSV.

From figure 9.2, it can also be seen that the trend of having registered NSV at the public setting has gone down significantly over the years in rural Ahmedabad same as urban Ahmedabad.

**Figure 9.1.**
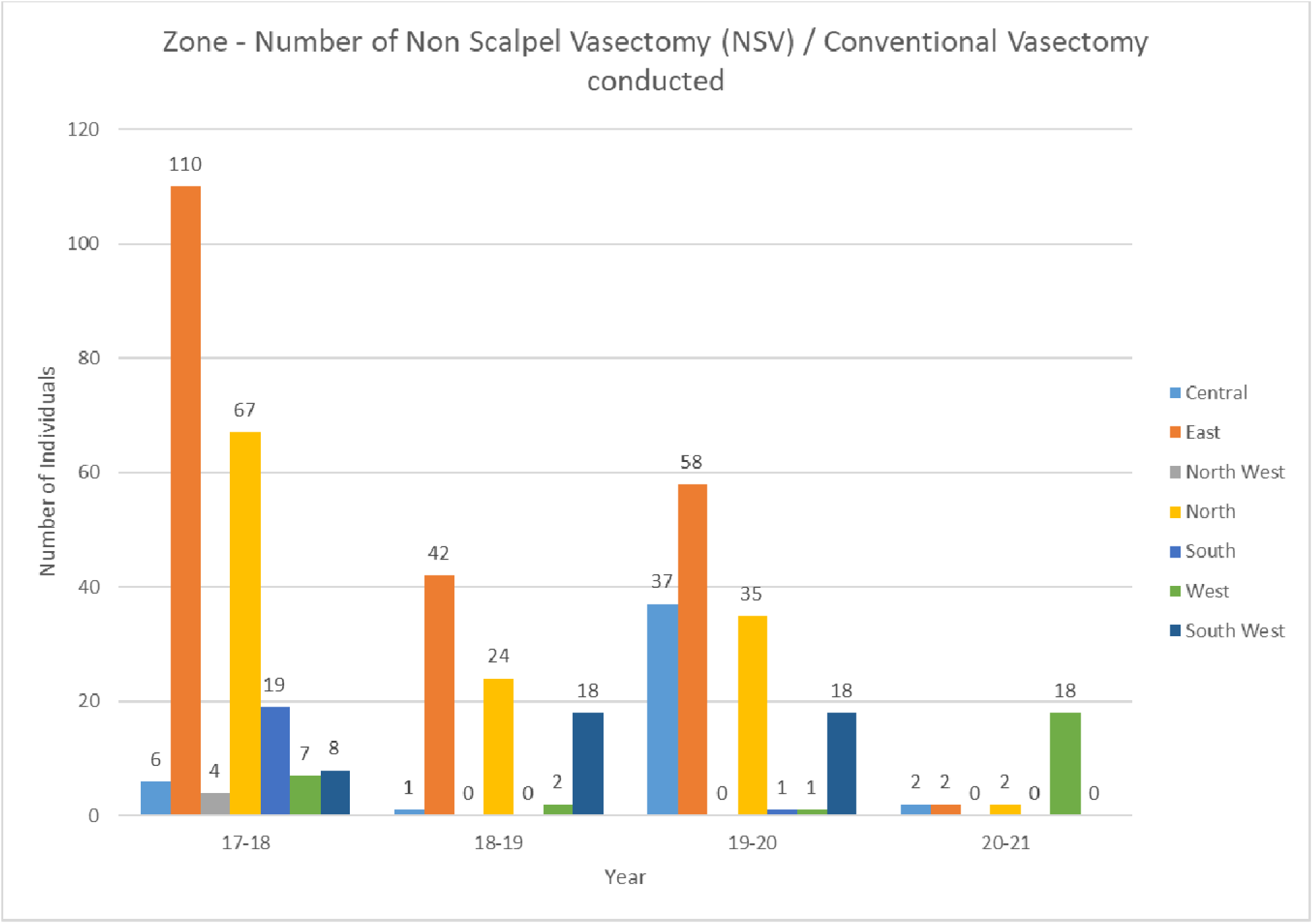

**Figure 9.2.**
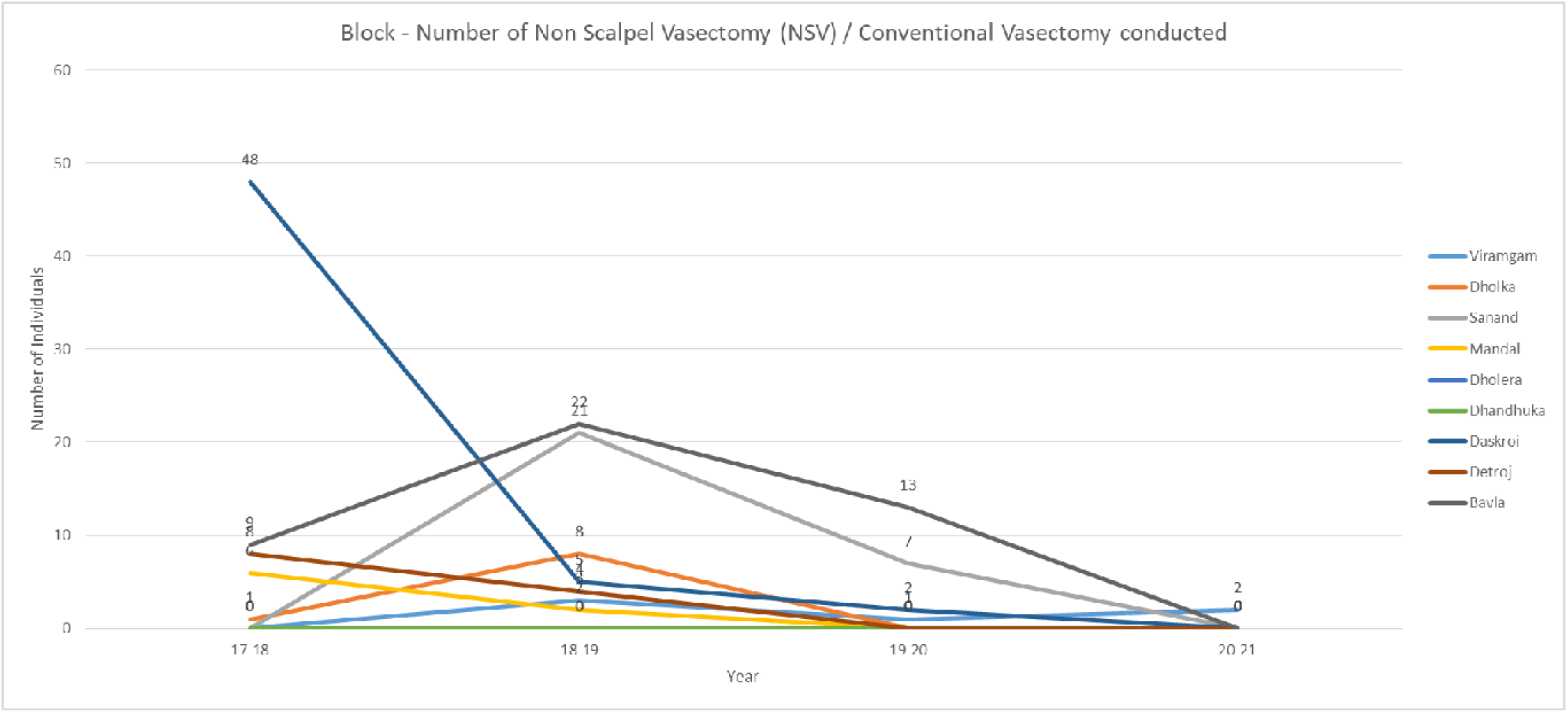

### 10) Female sterilization

Figure 10.1 shows us that the trend of having female sterilization remained similar with West zone showing the highest numbers in urban Ahmedabad has remained the same over the years except there is still an overall proportional decrease in female sterilization in all zones in 20-21.

Same as Figure 10.1, it can be seen in figure 10.2 that an almost constant number of procedures in all blocks in rural Ahmedabad takes a dip in 20-21, except Detroj.

**Figure 10.1.**
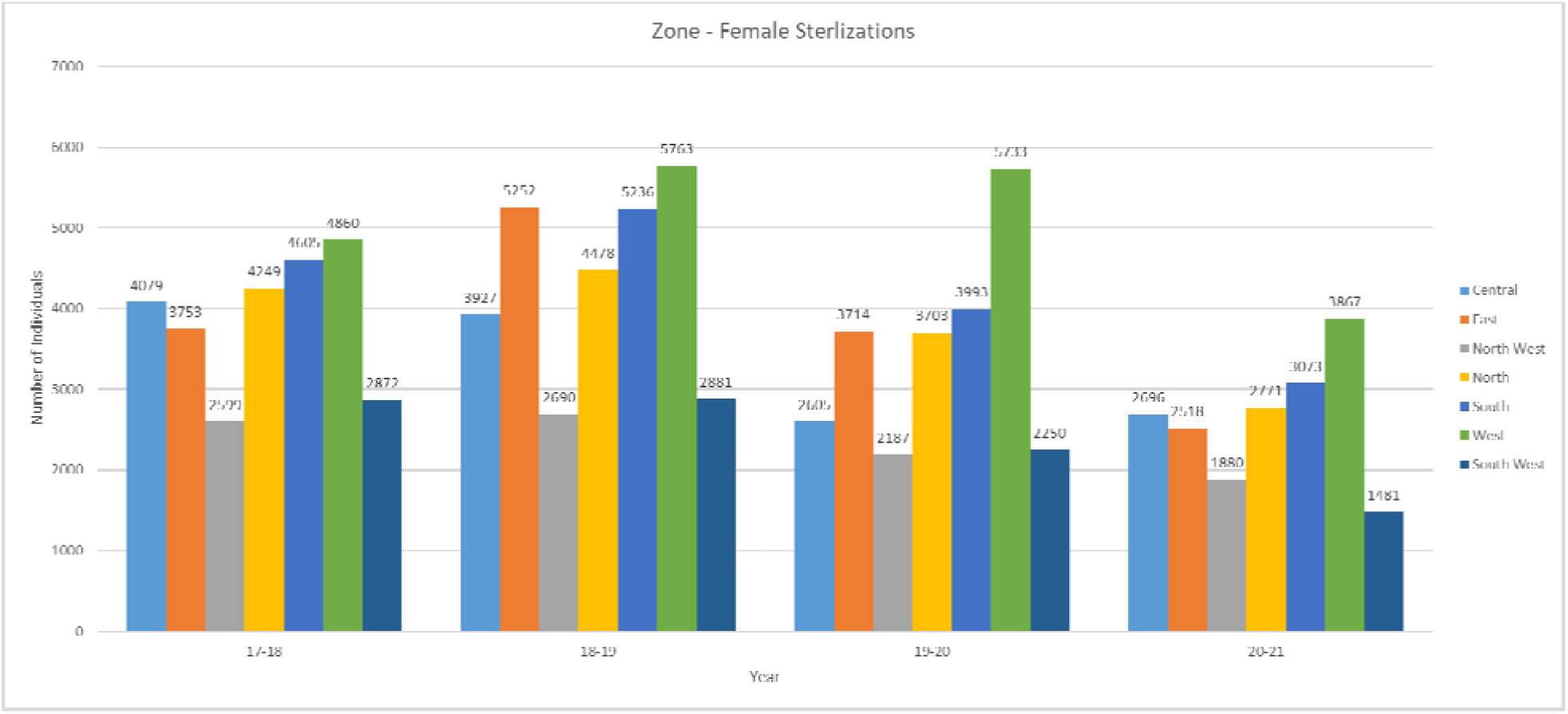

**Figure 10.2.**
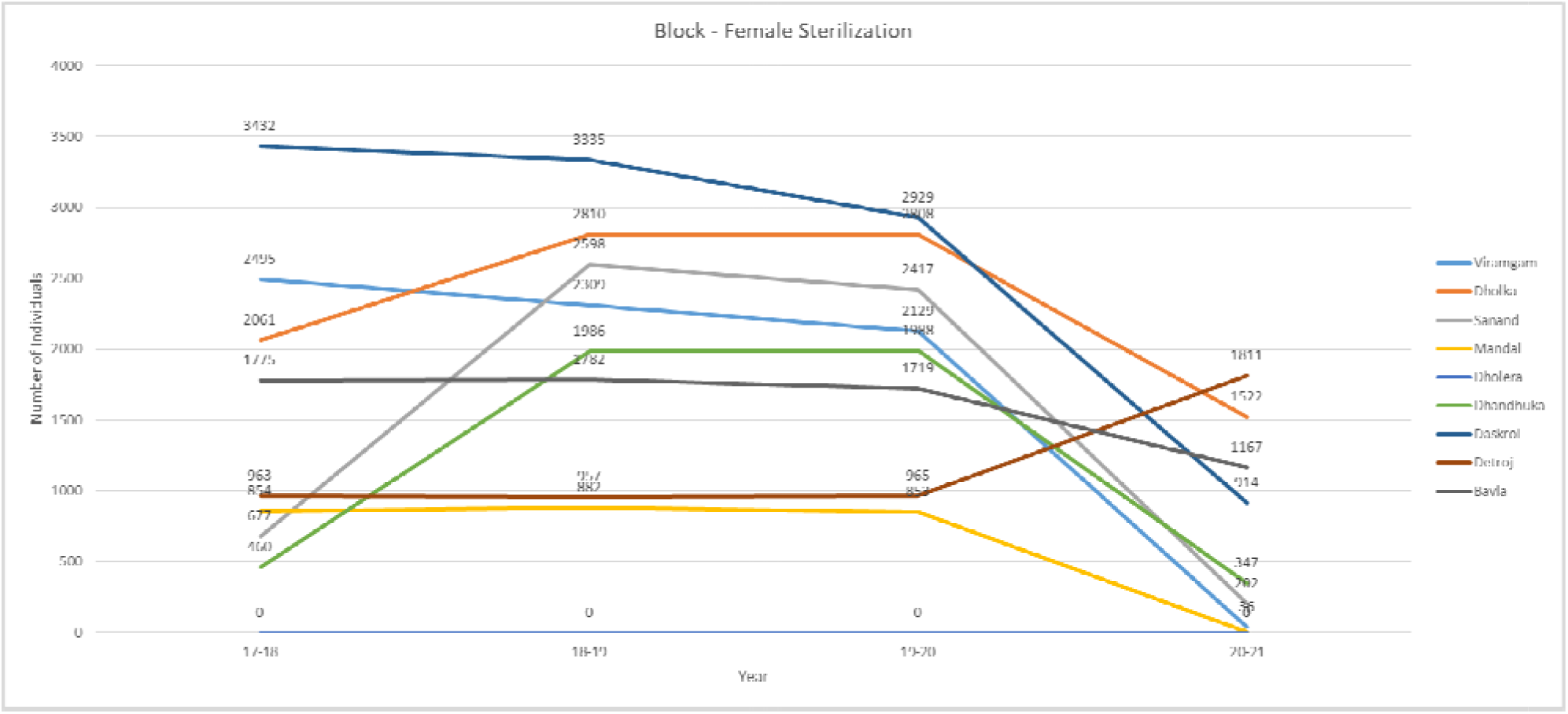

### 11) Children aged between 9 and 11 months fully immunized

A trend similar to Figure 1.1 can be seen in Figure 11.1 in the amount of children between age 9 to 11 months getting fully immunized, but it is not comparable with figure 7.1 live births meaning **registered pregnant women come to public setting to immunize their children but for delivery purposes, private hospitals are chosen over public settings. Suggesting the poor availability and facility in public settings when it comes to deliveries**.

Same as urban Ahmedabad, rural Ahmedabad in immunization status as per figure 11.2 is comparable with figure 1.2 registered pregnancies but not with figure 7.2 live births.

**Figure 11.1.**
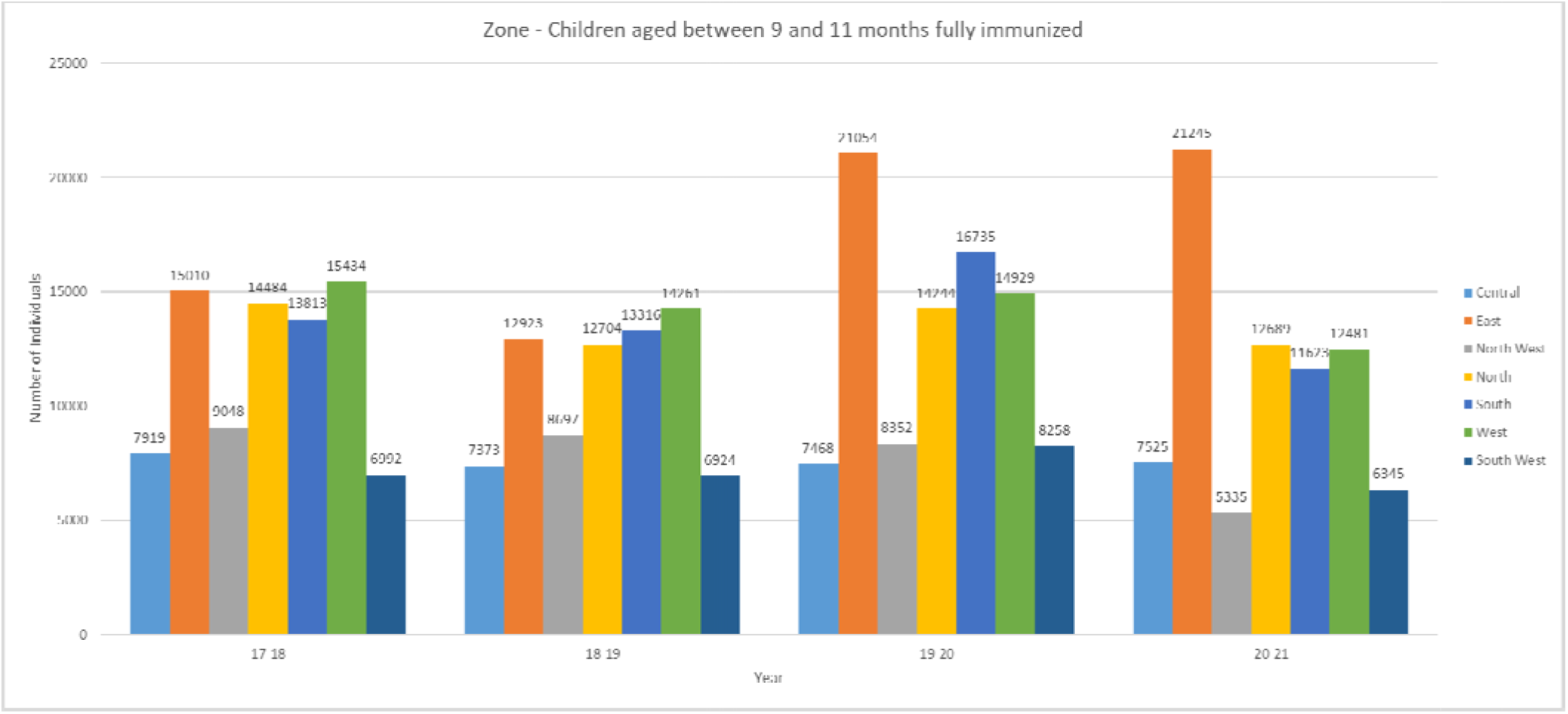

**Figure 11.2.**
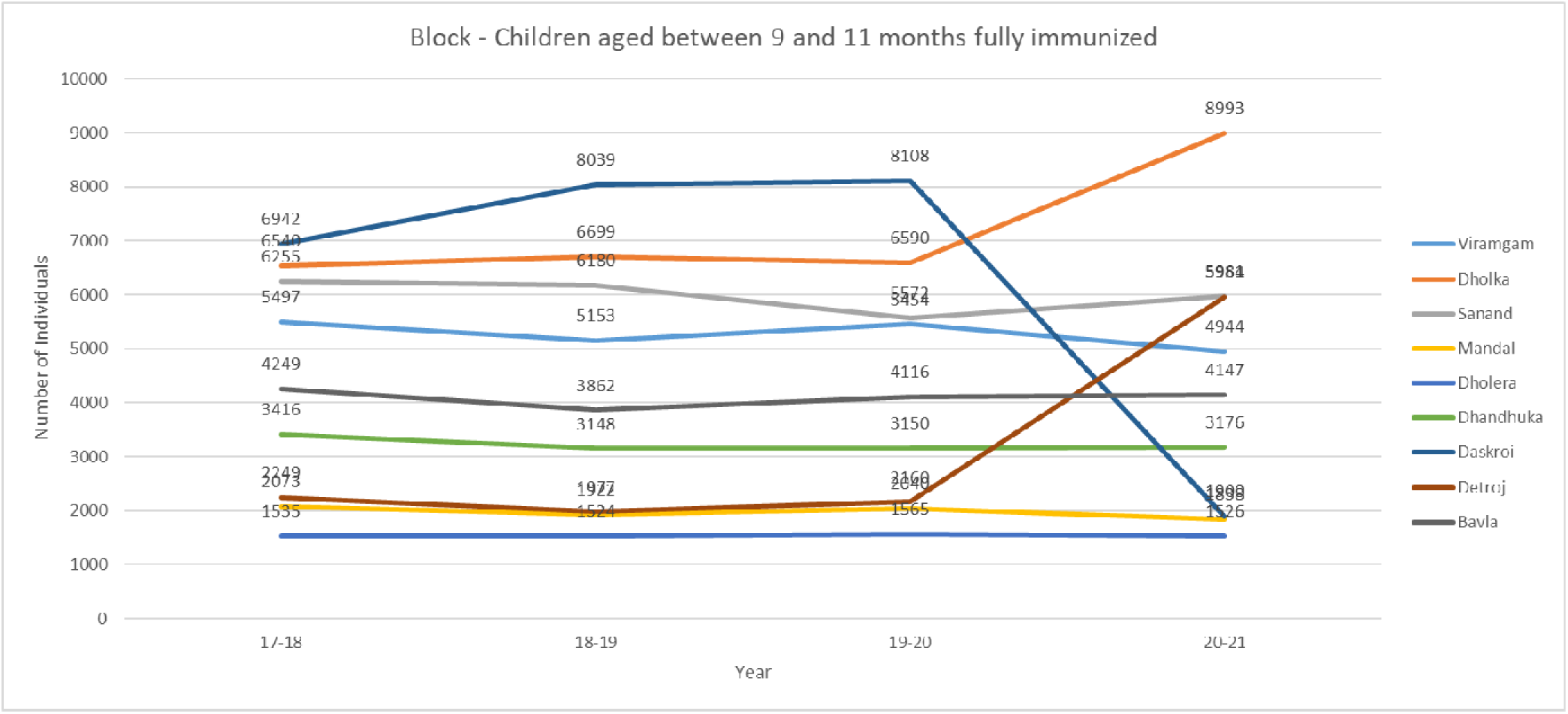

### 12) Number of severely underweight children provided Health Checkup (0-5 yrs.)

Figure 12.1 and 12.2 puts emphasis on the number of checkups provided to severely underweight children between age 0-5. Here in both Urban and Rural Ahmedabad, a common trend line can be seen showing a decrease in the years 20-21. **It is hard to understand whether the checkups were not provided or not needed from the data gathered.**

**Figure 12.1.**
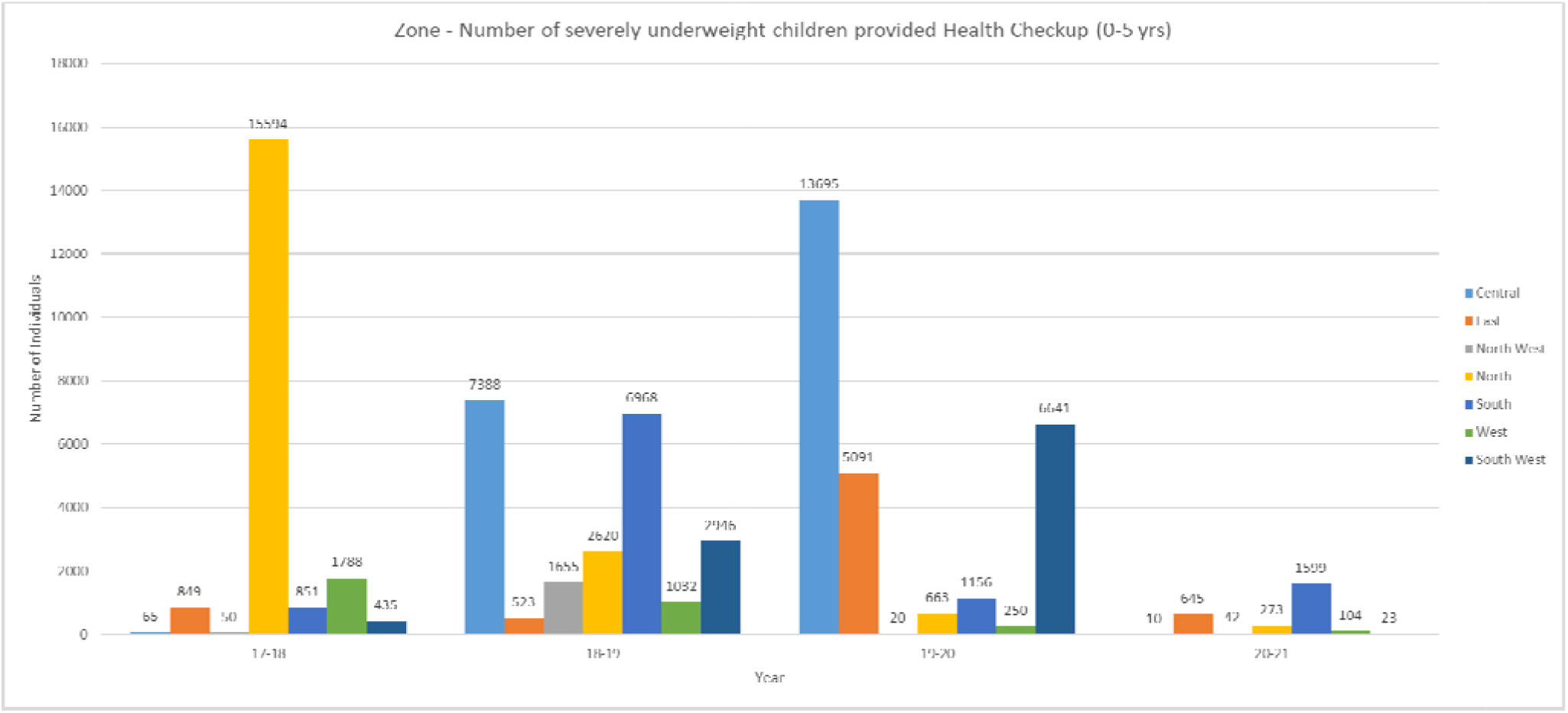

**Figure 12.2.**
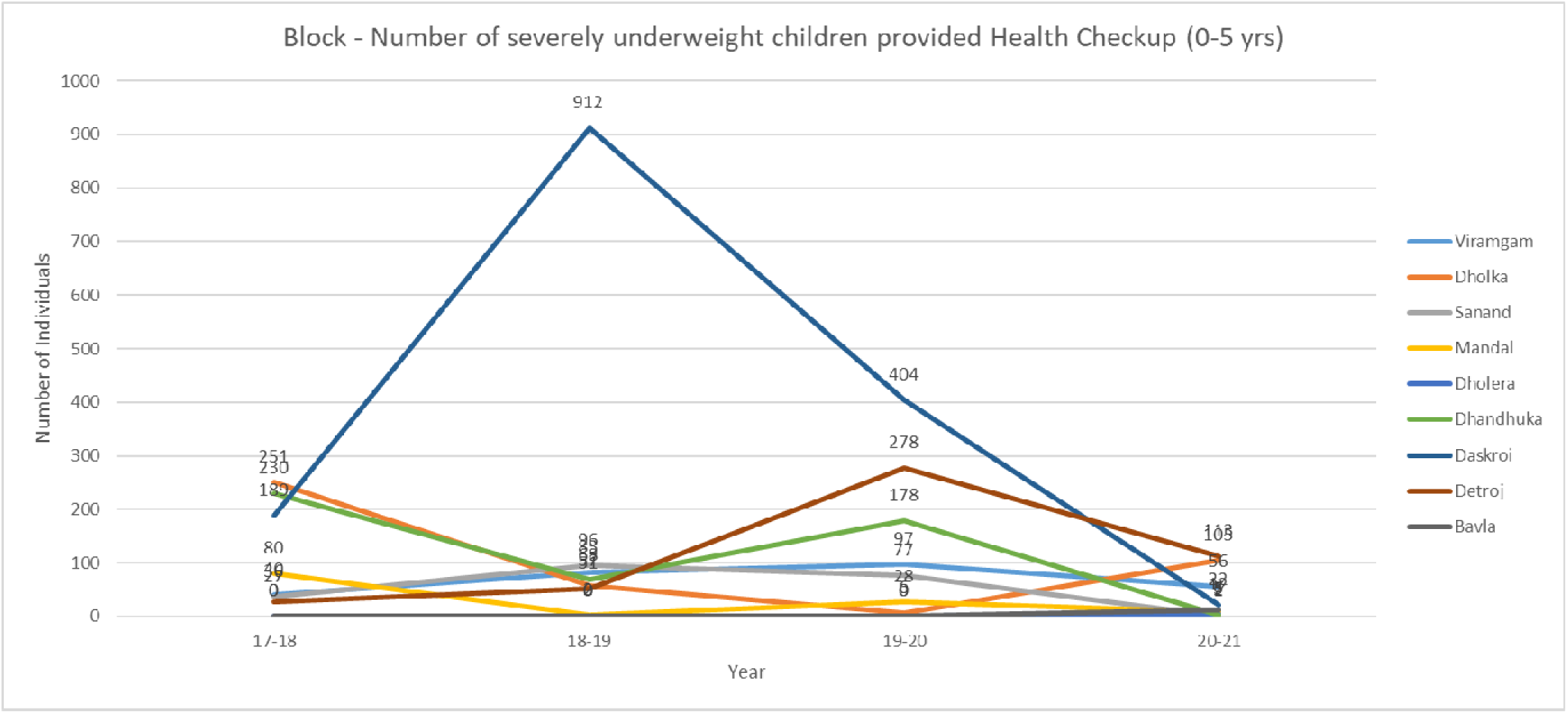

### 13) Childhood Diseases – Severe Acute Malnutrition (SAM)

Figure 13.1 and 13.2 same as figure 12.1 and 12.2 showing numbers of underweight children showing diagnosed SAM and a similar trend in both urban and rural is seen ending as a sudden drop in 20-21 **not clearing whether there were actually less cases or less cases were diagnosed.**

**Figure 13.1.**
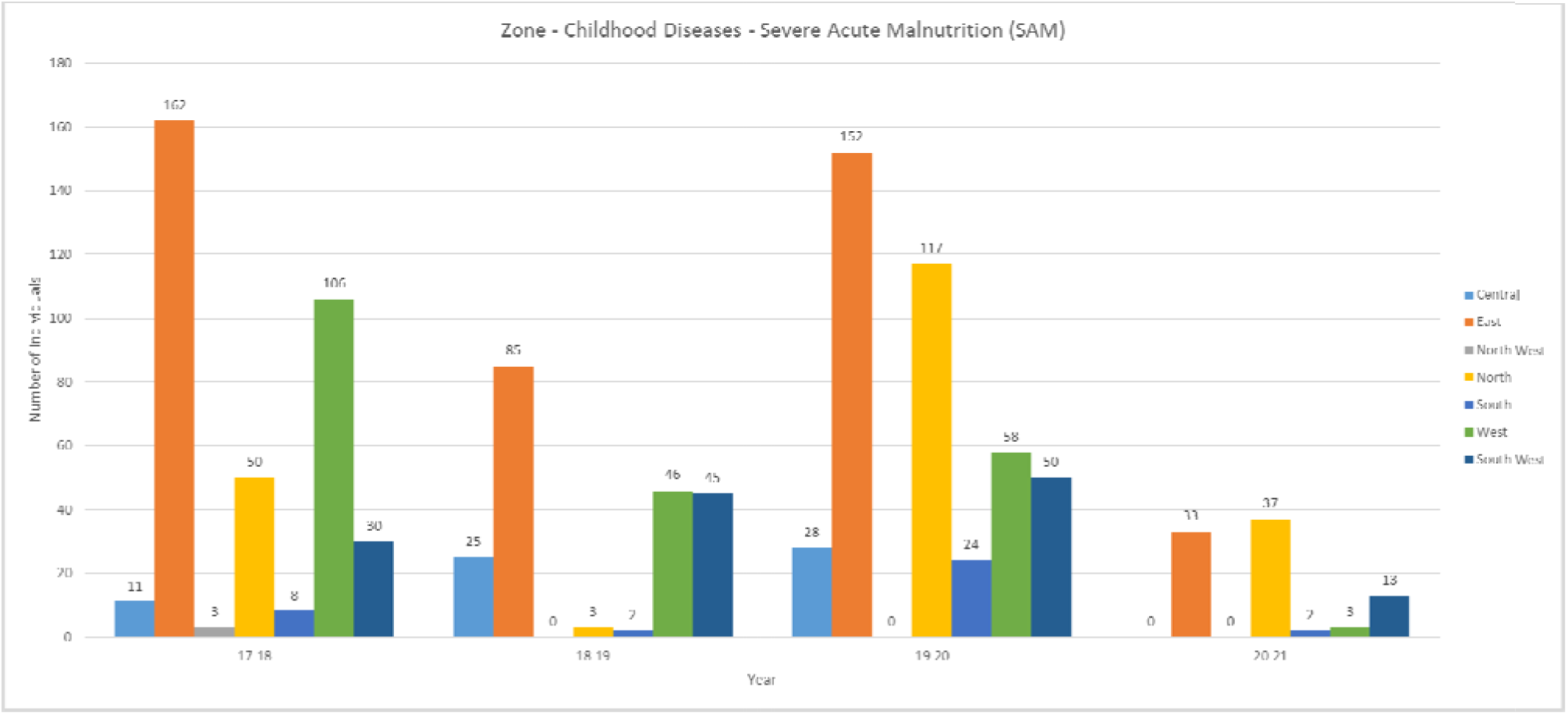

**Figure 13.2.**
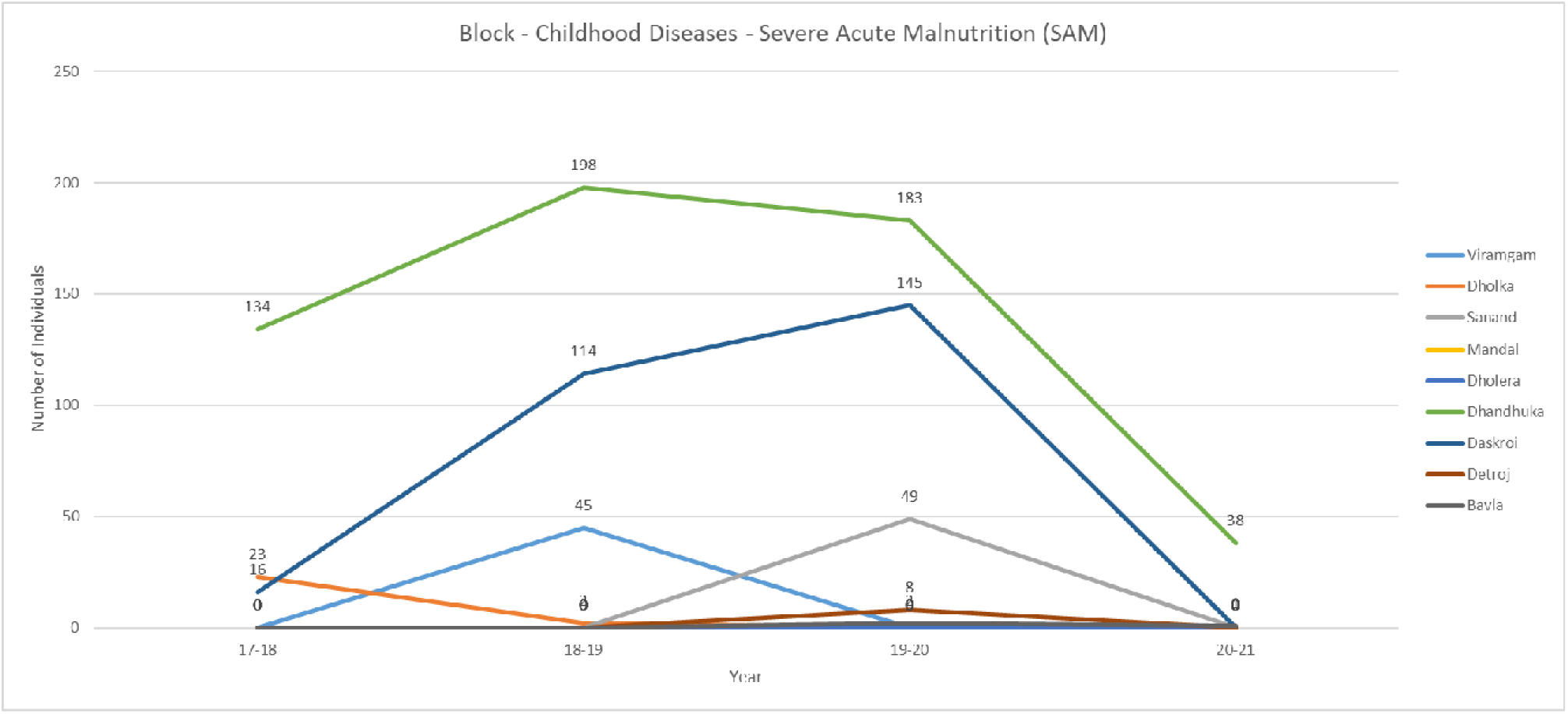

### 14) Infant deaths within 24 hours (1 to 23 hours) of birth

Figure 14.1 and 14.2 demonstrate here the Infant mortality has decreased significantly over the years in both Urban and rural Ahmedabad. The numbers shown here are absolute numbers and no ratio can’t be calculated as the number of live births reported is faulty as seen in figure 7.1, 7.2 and discussion 1.

**Figure 14.1.**
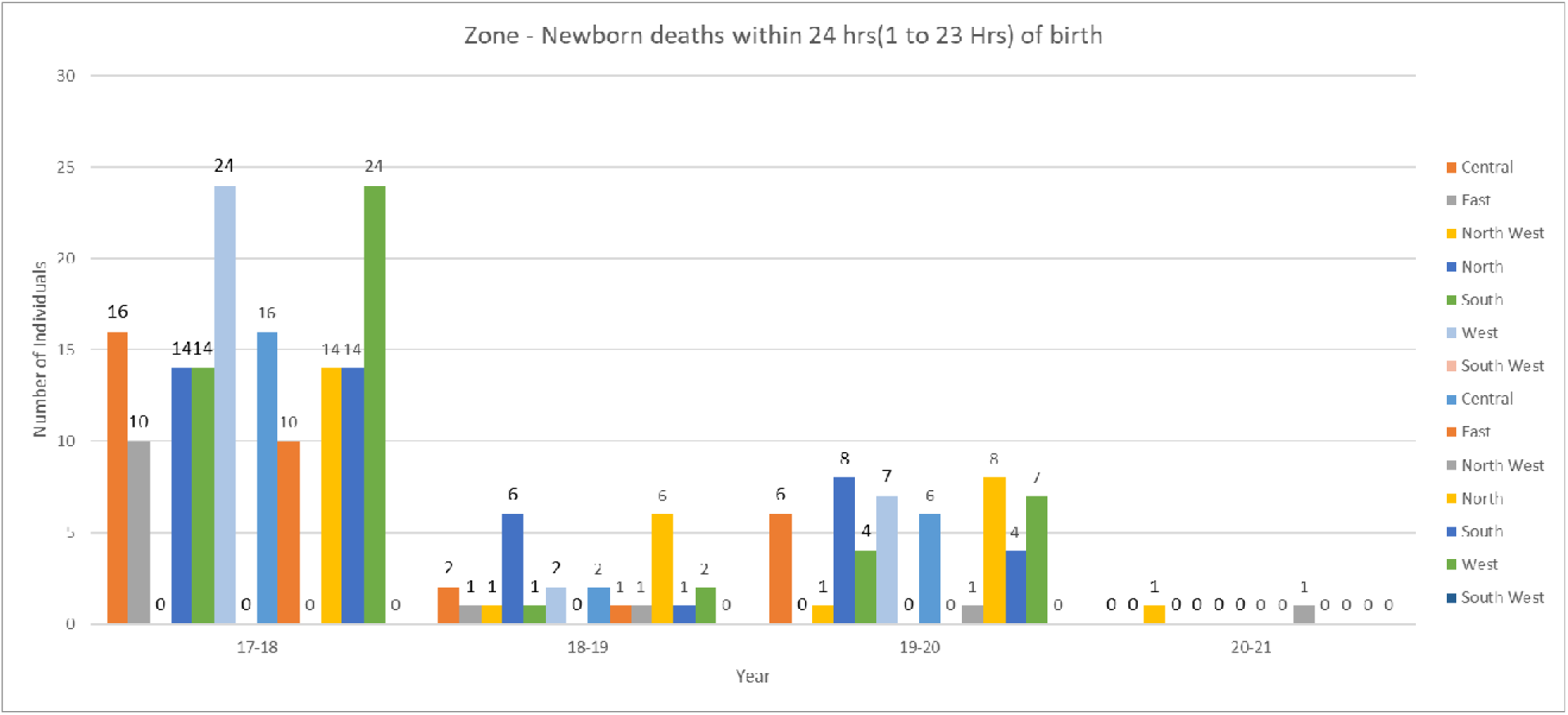

**Figure 14.2.**
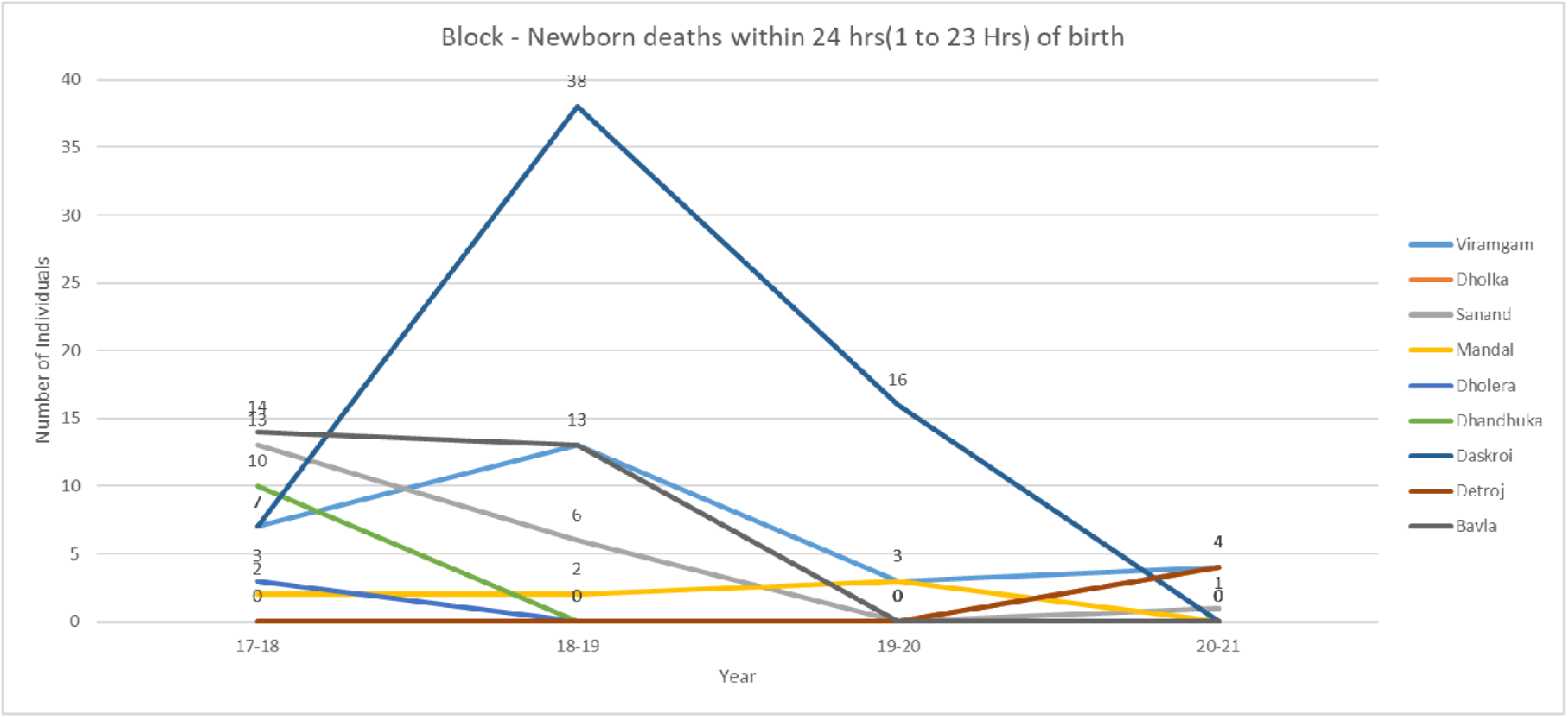

### 15) Neonatal Deaths (1 day to 28 days)

Figure 15.1 shows high amounts of neonatal mortality in rural Ahmedabad in the years 17-20. These deaths are mainly due to the large number of deaths occurring in Dholka block. Otherwise in other blocks, the numbers are low.

Urban Ahmedabad shows a similar trend that can be seen in NFHS-5 Gujarat and NFHS-4 India but mortality in 20-21 is very low which cannot be understood.

**Figure 15.1.**
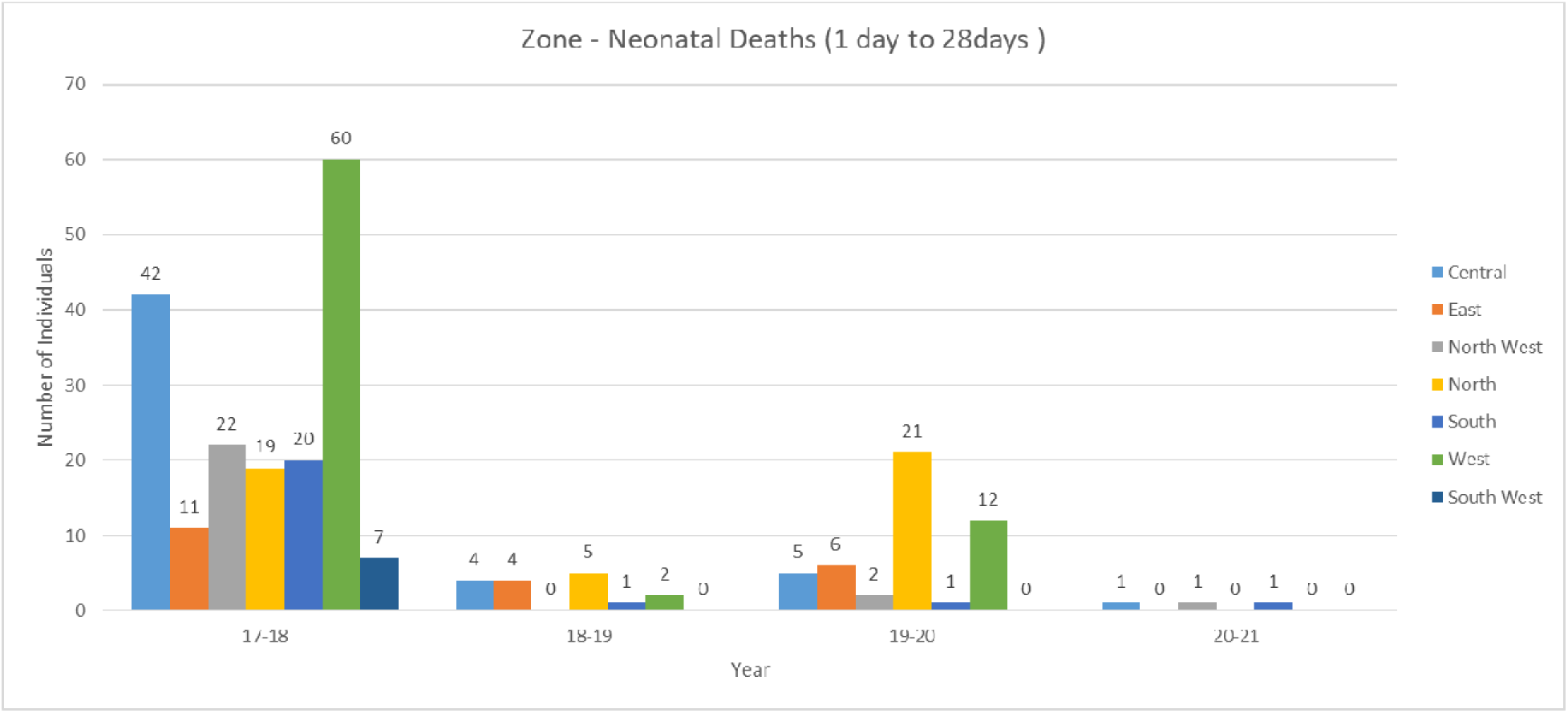

**Figure 15.2.**
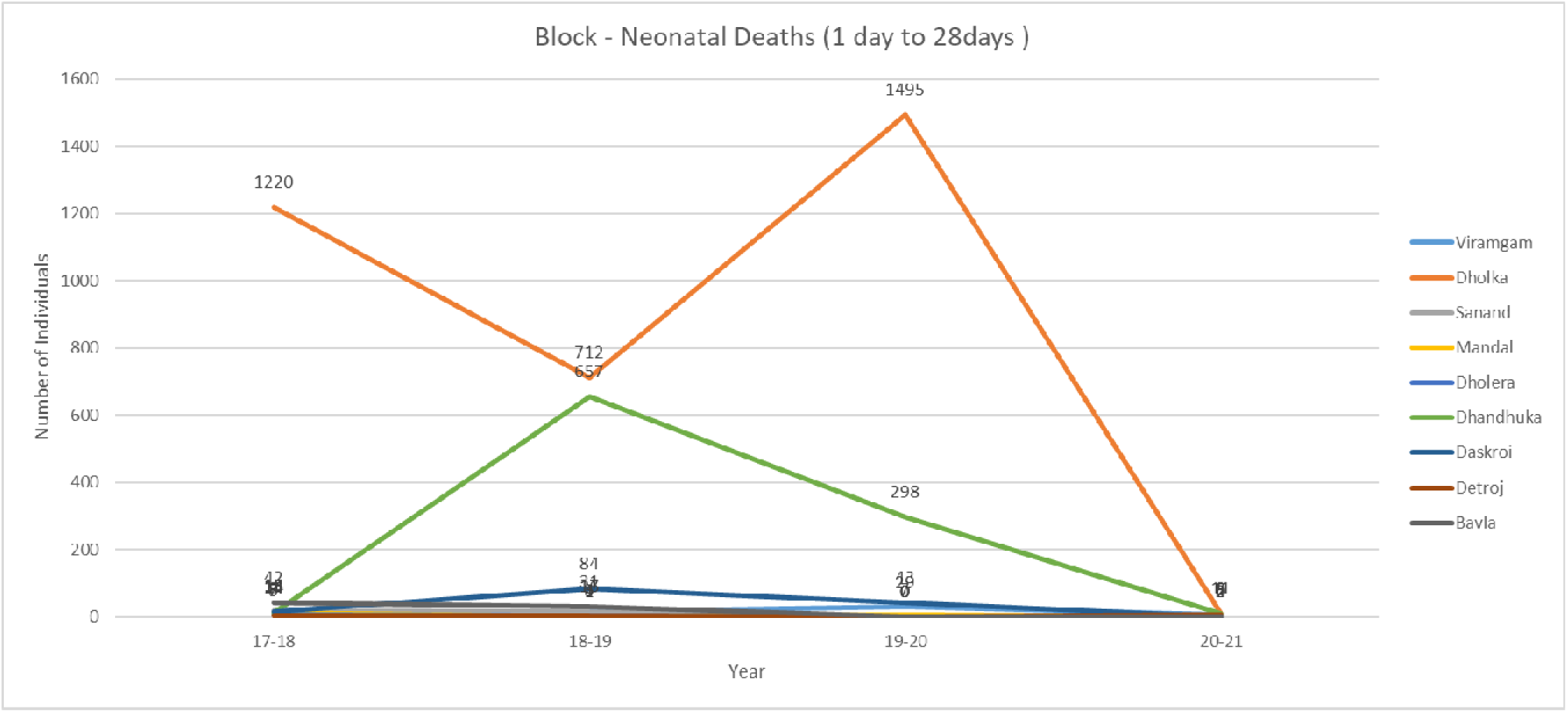

**Figure 15.3.**
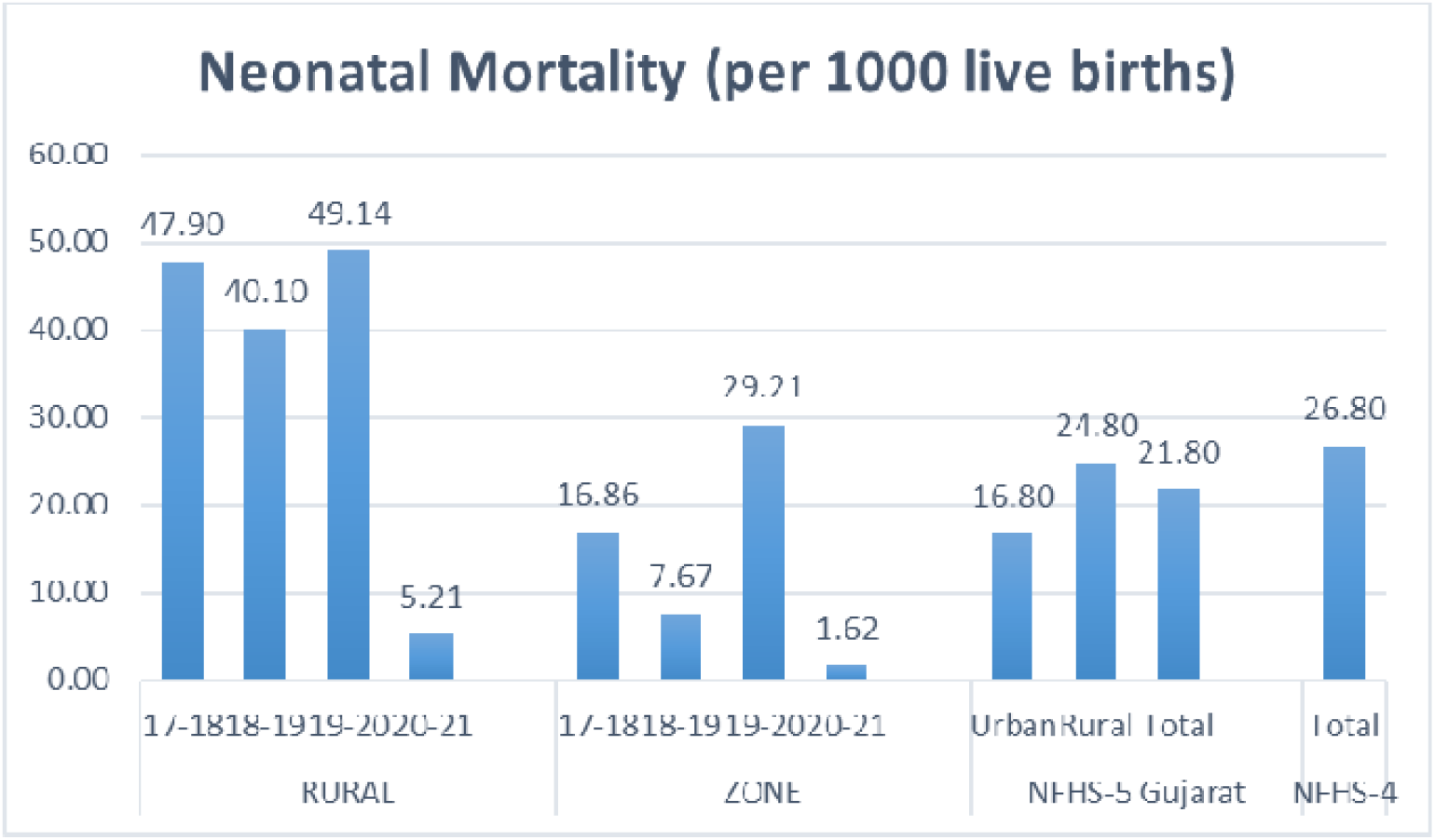

### 16) Infant deaths (1 month to 1 year)

Figure 16.1 denotes that Rural and Urban Ahmedabad has been able to control infant mortality with great success as NFHS-5 Gujarat numbers are high but NFHS-4 India numbers are even higher.

**Figure 16.1.**
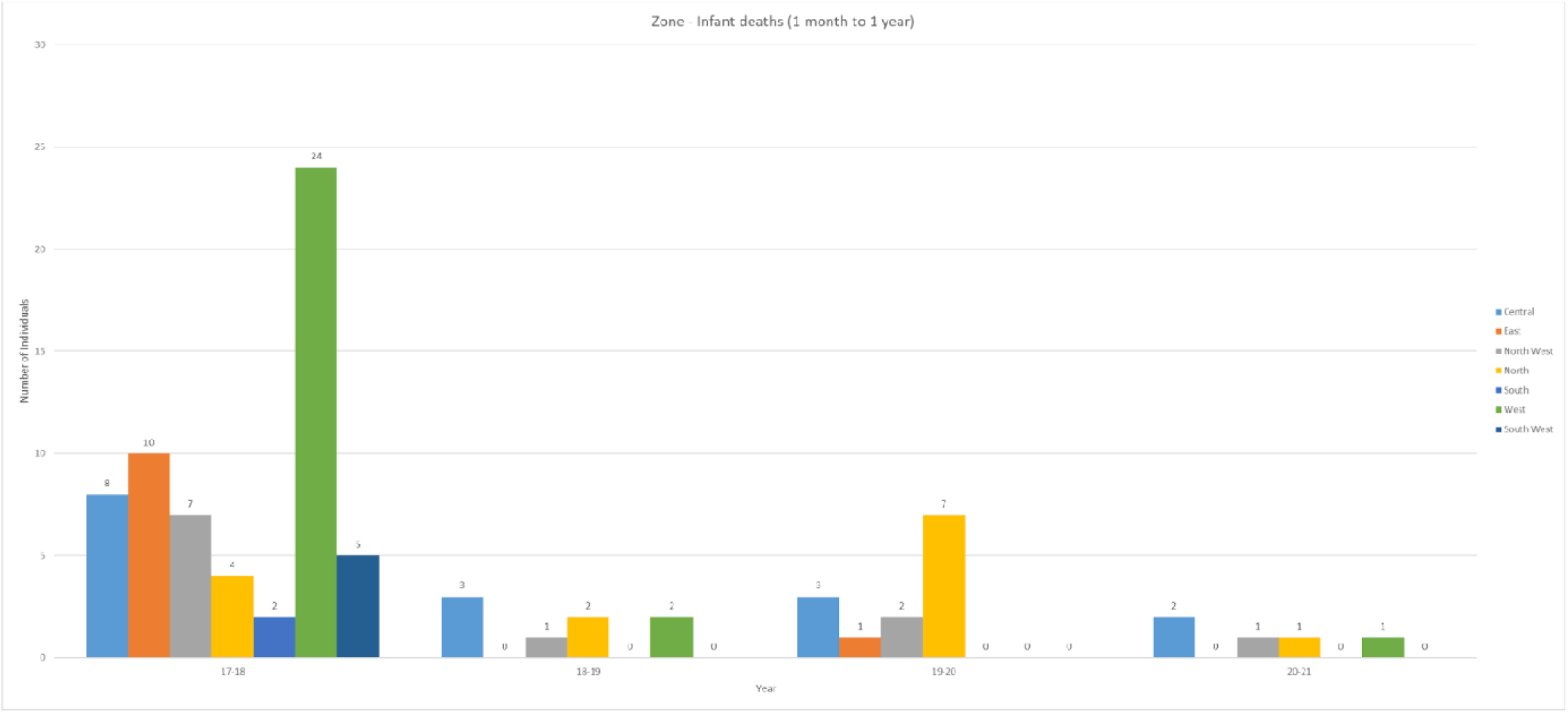

**Figure 16.2.**
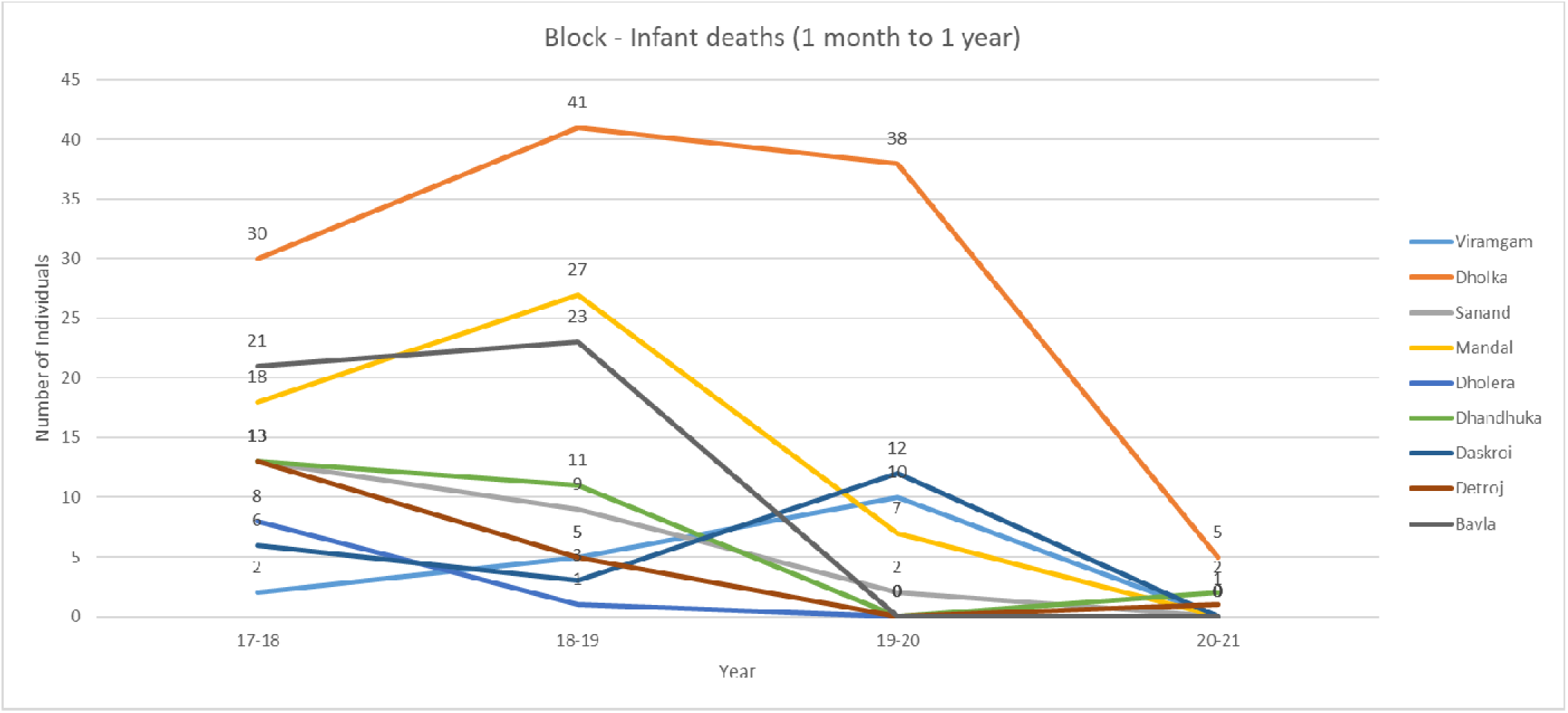

**Figure 16.3.**
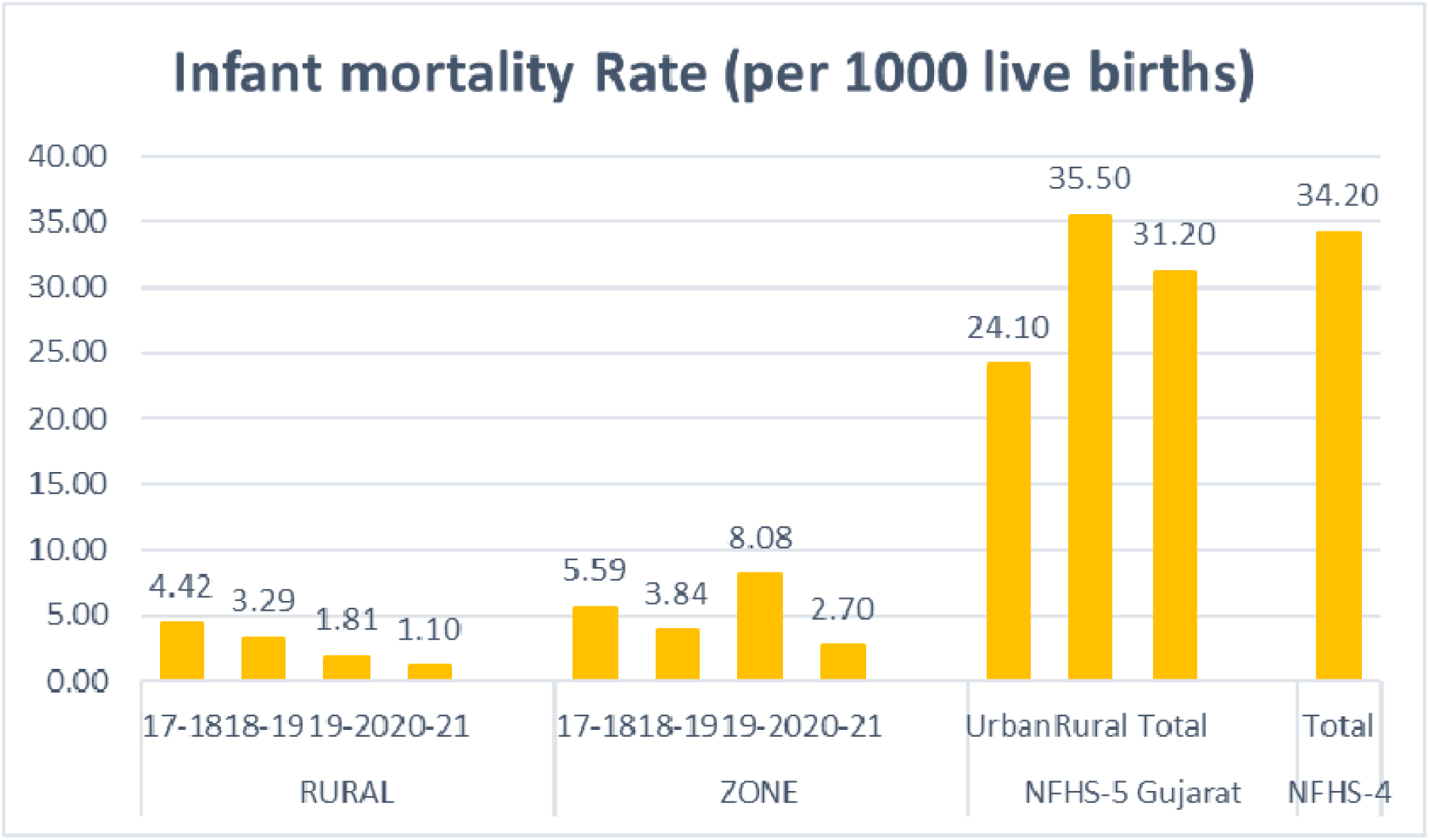

### 17) Under 5 deaths (1 year to 5 year)

Figure 17.1 denotes that Rural and Urban Ahmedabad has been able to control Under 5 mortality with great success as NFHS-5 Gujarat numbers are high but NFHS-4 India numbers are even higher.

**Figure 17.1.**
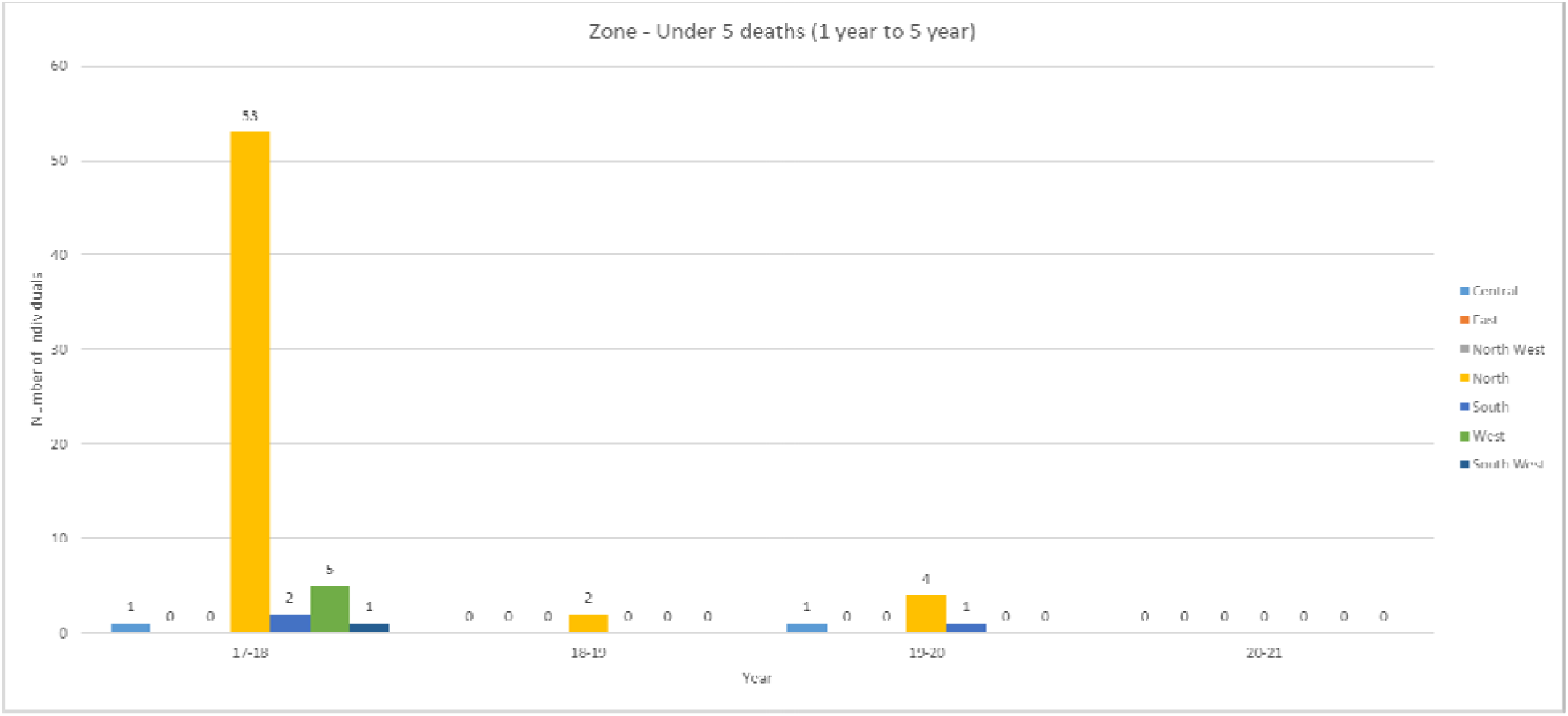

**Figure 17.2.**
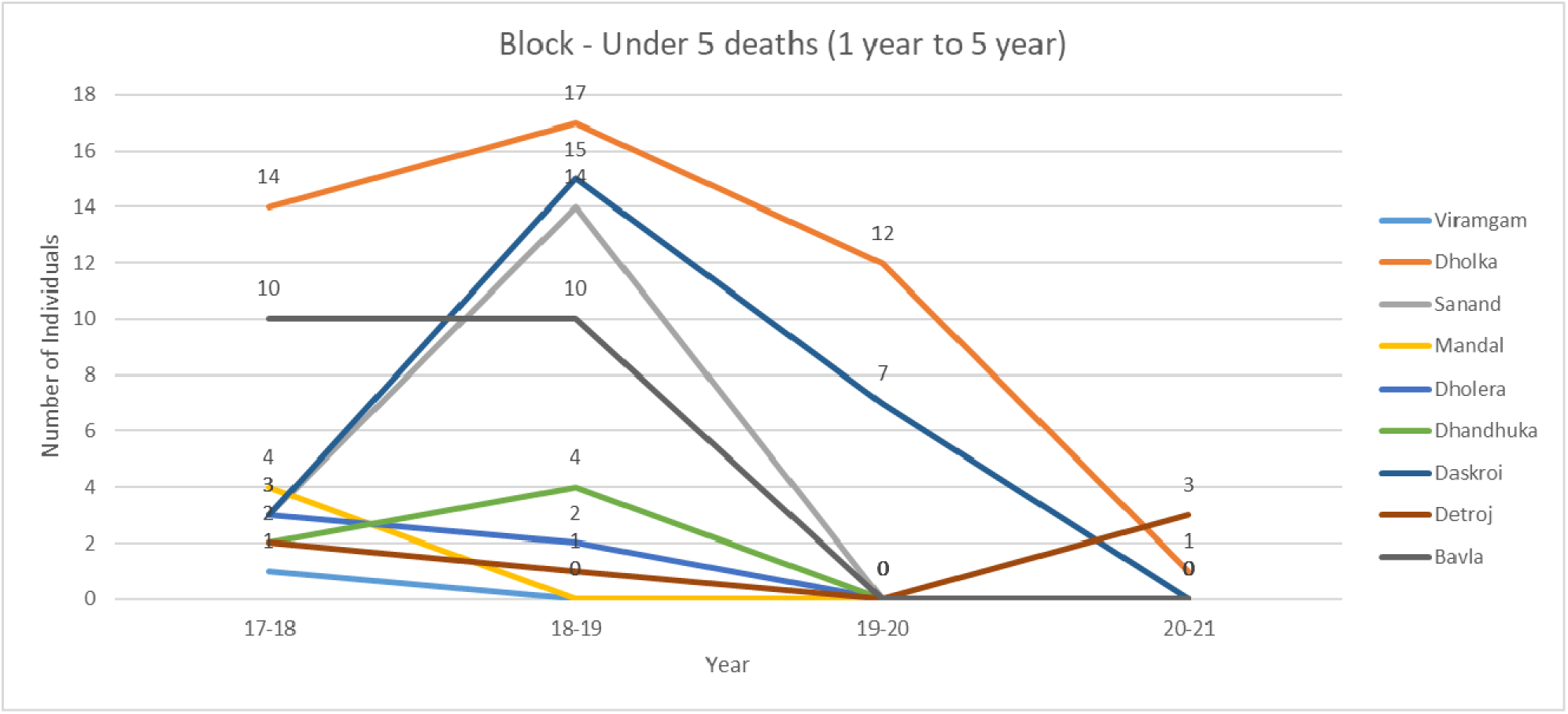

**Figure 17.3.**
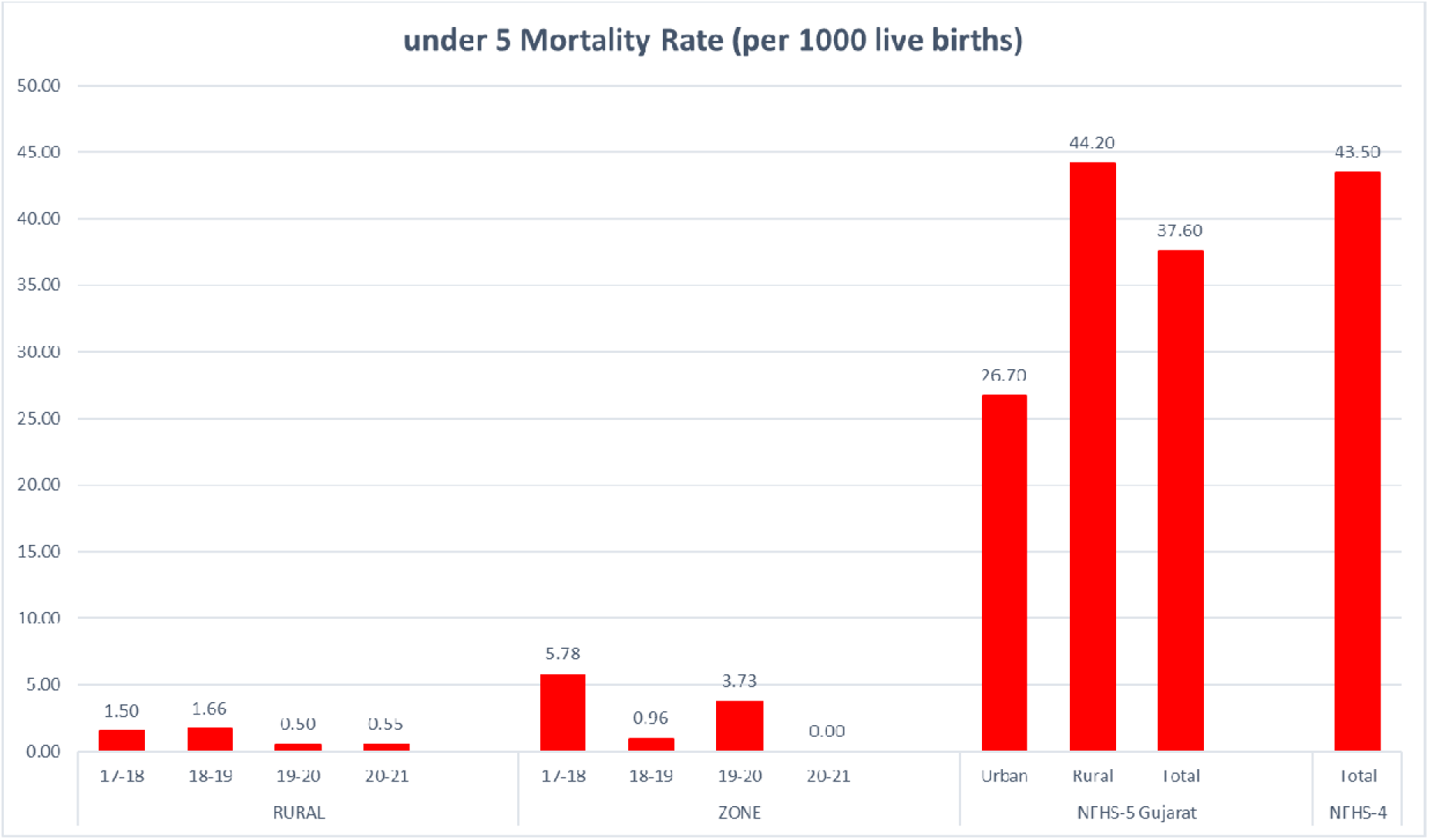

### 18) Maternal Deaths

Figure 18.1 and 18.2 shows a small amount of maternal death numbers but these are absolute numbers. Also, we can’t compare this with NFHS data as NFHS doesn’t give Maternal Death ratio or numbers.

**Figure 18.1.**
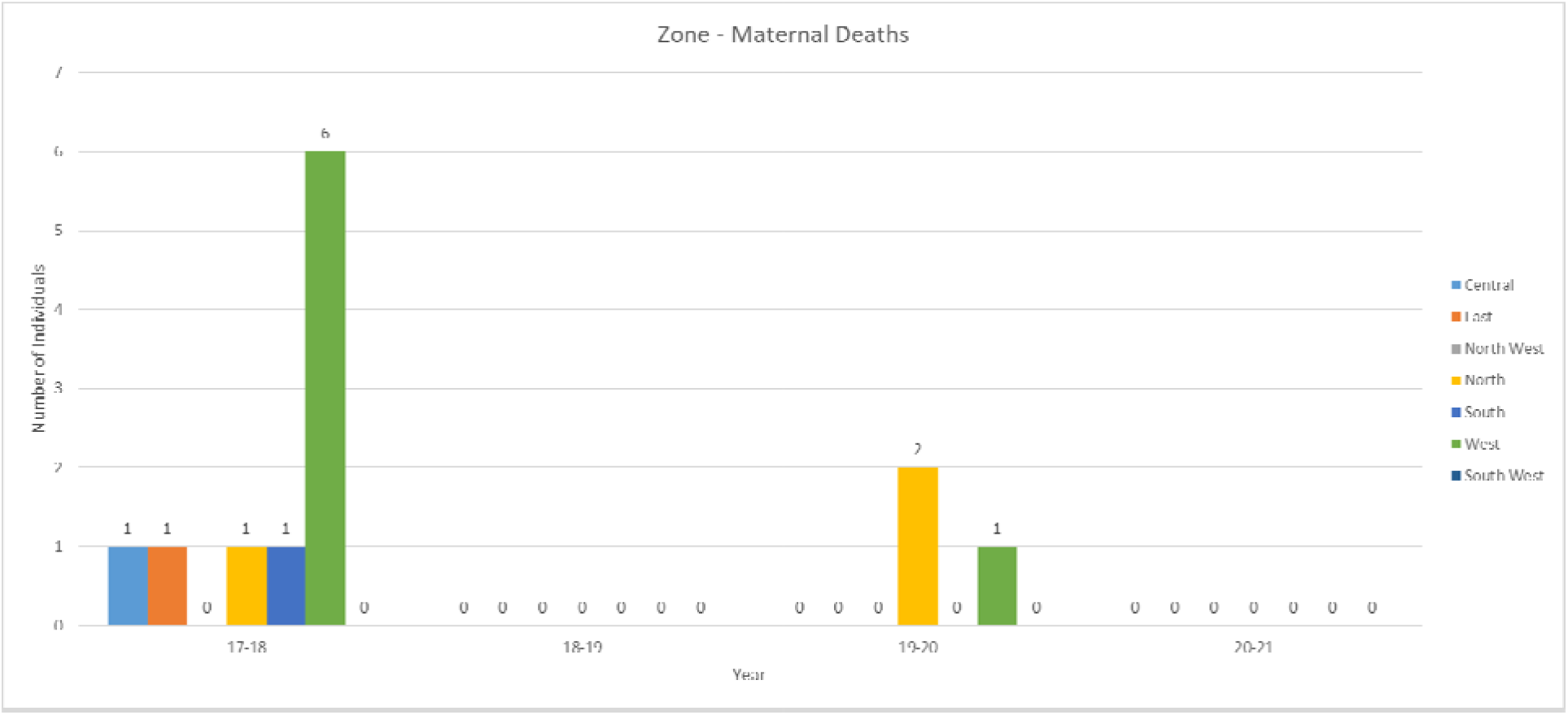

**Figure 18.2.**
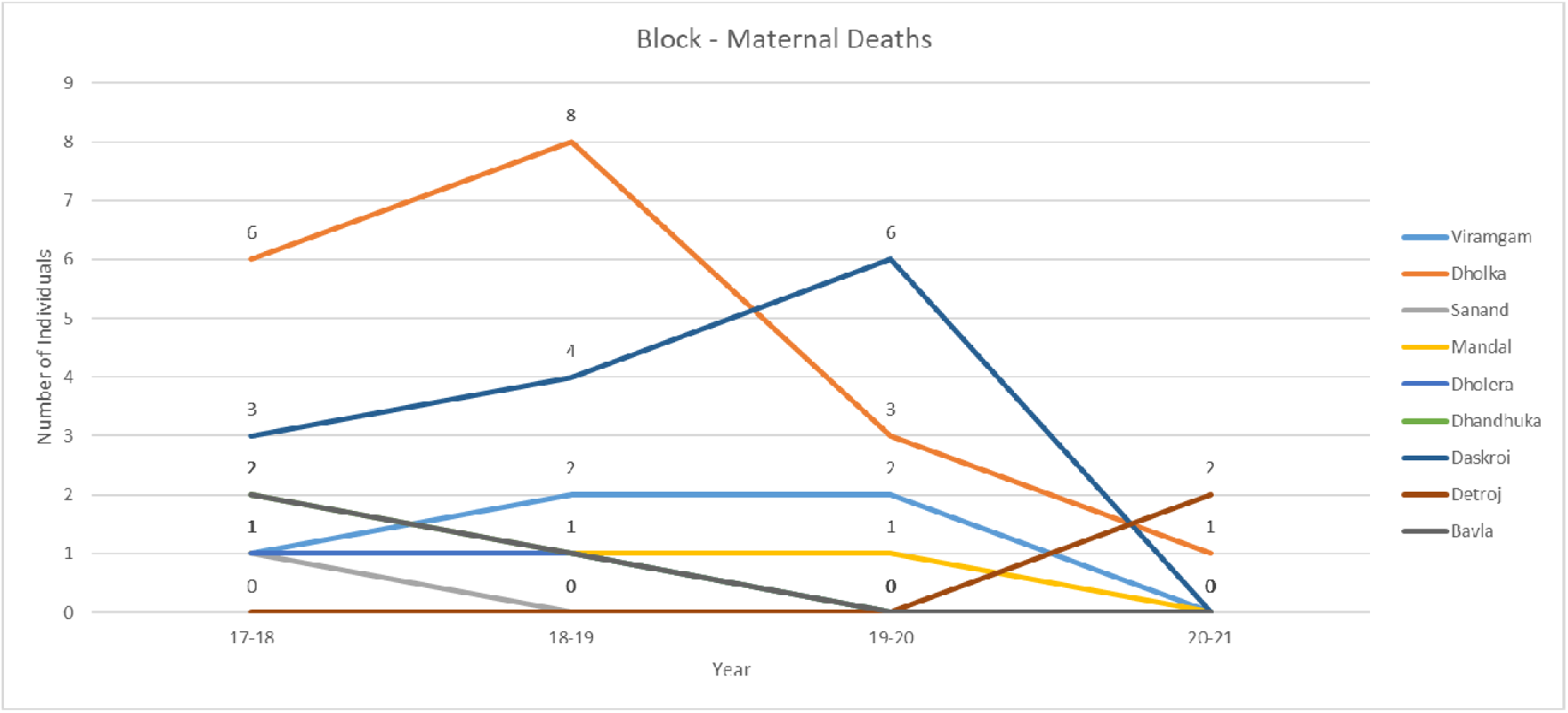

## Conclusion

This study generates valuable information which can be utilized by local governing bodies to improvise the services given by these health centers. In this study, it was particularly found that Data Management at various levels is faulty and there are profound misconducts in Data Collection, Data Gathering, Data Analysis and Data preserving. There is a visible difference between how Rural data is managed versus how Urban data is managed. Results found with available data in Ahmedabad Urban or Ahmedabad rural seemed better in regards to Reproductive and child health care when compared to average of the entire Gujarat state and India. Better methodology and techniques are now a necessity for quality Data Recording, Management and Safe-keeping.

## Summary

This study paves the road for future analysis of the services to improve the indicators.

This study was conducted to find out a pattern and efficiency of determining the quality of a single PHCs work, work being done in a Zone or a block, and work being done for Reproductive and Child health in entire district. Data’s from all individual UHCs from Ahmedabad city areas and data from all PHCs from Ahmedabad district was gathered and pooled to find out totals for Zones and Blocks and those were compared with National averages and state averages to assess quality of work being Carried out in Zone and Blocks.

There is ample data available in PHCS and UPHCs but that data is not utilized and analyzed properly for the betterment of the community. This study actually demonstrates what should have been done for years and what should be done now in terms of resources and infrastructure and whether its re-distribution is needed or not. We hope to trigger re-evaluation of working processes and make progress at a sub-center grass root level so that we can re-think about what we lack in our goals and in UHC2030 program aim and work to achieve it faster and more efficiently.

Data from private settings and hospitals are not covered and accounted for which leads to large irregular variations which is not possible in reality. Hence, the data collected should be inclusive of the private hospitals also when it comes to RCH. The data present at UHCs and PHCS level and the data shown to the world has no connection and are grossly mismatched. There are some portions in the report too which cannot be explained with a logical hypothesis. There seems to be gross misconduct and mishandling of data knowingly or unknowingly which is not being corrected.

## Data Availability

All data produced in the present study are available upon reasonable request to the authors

